# The Efficacy of transcranial Direct Current Stimulation (tDCS) on Emotional Processing

**DOI:** 10.1101/2024.10.09.24315160

**Authors:** Fatemeh Akbari, Abdolvahed Narmashiri

## Abstract

While some research has reported the positive impacts of transcranial direct current stimulation (tDCS) on emotional processing, the conflicting results and variations in study quality and design make it challenging to draw firm conclusions about its effects. To address this issue, we conducted a meta-analysis of the published literature on tDCS effects on emotional processing. We evaluated the effects of anodal and cathodal tDCS on emotional processing by analyzing a total of 32 articles and a combined sample of 2,107 participants. Our study illustrates the significant effects of both anodal and cathodal tDCS on emotional processing. The results highlight significant differences in how tDCS stimulation parameters influence emotional processing, demonstrating that frontal tDCS enhances emotional outcomes more effectively than stimulation of other brain regions. Key findings include that longer stimulation durations (over 20 minutes) and higher current intensities (2 mA) yield better results, with online tDCS being more effective than offline stimulation, particularly in younger participants and in non-clinical populations. Our results reveal that tDCS can effectively enhance emotional processing, offering valuable insights into the potential benefits of this method for emotion improvement.

## 1. Introduction

Transcranial direct current stimulation (tDCS) is a non-invasive technique that delivers a mild electrical current to the scalp, which can influence cortical excitability by adjusting the resting membrane potential of neurons. The effect varies with electrode polarity: anodal stimulation tends to enhance neuronal excitability, while cathodal stimulation generally reduces it. This current flow is believed to modulate cortical excitability in the targeted region (Jamil and Nitsche 2017). As a result, tDCS serves as a useful tool for exploring the brain regions and networks associated with particular cognitive functions (Narmashiri and Akbari 2023), and can also be applied to improve cognitive performance such as emotional processing (Gupta and Mittal 2021, Estaji, Hosseinzadeh et al. 2024).

Emotional processing is a broad term referring to various mental processes from emotional regulation (Clarke, Van Bockstaele et al. 2020) to emotional valence (La Malva, Di Crosta et al. 2024). Research has demonstrated that anodal tDCS, which increases excitability, applied over the left dorsolateral prefrontal cortex (lDLPFC), along with cathodal tDCS, which decreases excitability, over the right DLPFC, can enhance positive emotion processing and reduce the impact of negative emotions (La Malva, Di Crosta et al. 2024). Furthermore, tDCS has been shown to improve emotion regulation in both healthy individuals and those with mood disorders. For instance, anodal tDCS over the prefrontal cortex has been found to bolster cognitive control over emotional distractions, which can be especially beneficial for individuals experiencing depression (Zakibakhsh, Basharpoor et al. 2024). Additionally, high-definition tDCS (HD-tDCS) targeting the primary somatosensory cortex (S1) has been linked to enhanced interoceptive abilities, critical for emotional awareness and processing (Nitsche, Koschack et al. 2012). Repeated sessions of prefrontal tDCS have also been associated with reduced psychological distress and improved mental health outcomes, including better quality of life and sleep quality (Plewnia, Schroeder et al. 2015). While some studies have reported positive effects of tDCS on emotional processing, others have noted no significant impact, highlighting variability in outcomes across different investigations, likely due to differences in experimental design and measurement methods. Moreover, considerable methodological variations between experiments contribute to inconsistent findings. Recent studies on emotional processing have pooled results from experiments with differing tDCS parameters—such as stimulation site, current intensity, duration, total sessions, online vs. offline application, outcome measures, and participant types—to assess the reliability of tDCS effects. While some research has reported positive impacts of tDCS on emotional processing, the conflicting results and variations in study quality and design make it challenging to draw firm conclusions about its effects on emotional processing.

To address this issue, we conducted a meta-analysis to evaluate the effects of tDCS on emotional processing. Our analysis pooled studies with varying stimulation parameters, methodologies, and other factors such as stimulation site, current intensity, duration, total stimulation time, online versus offline stimulation, outcome measures, and population characteristics. The study had two primary objectives: first, to determine the effects of anodal and cathodal stimulation on emotional processing, and second, to investigate how tDCS stimulation parameters influence emotional outcomes.

## 2. Methods

### 2.1 Search strategy

A comprehensive literature search was conducted using PubMed, Scopus, and Web of Science to identify related studies published from 1993 to October 2021. The search strategy consisted of two keywords: “intervention” and “emotion”, with the former referring to tDCS and related terms. Literature searches were restricted to studies published in English and involving human participants. All search results were combined and duplicates were eliminated.

### 2.2 Inclusion criteria

The following criteria were used to select articles for inclusion in this meta-analysis: (I) human subjects, (II) use of tDCS as an intervention, (III) reported quantitative results for both the tDCS and sham groups, with differences between the groups also clearly reported, (IV) sample size and mean (SD) data provided as required for standard meta-analysis, (V) publication in English, and (VII) peer-review reviewed.

### 2.3 Exclusion criteria

In this meta-analysis, four following criteria for exclusion of articles were used: I) articles that used different ways in combination with tDCS, II) studies without a sham group, III) duplicate publications, IV) review and conference studies

### 2.4 Study selection and data extraction

Databases were searched, and descriptions of all studies matching the search terms were recorded. These studies were then evaluated to determine whether they satisfied the predetermined inclusion and exclusion criteria. The following items were extracted for selected studies: (I) general data, such as the year of publication, the author’s name, and whether emotions were reported; (II) individual data, such as the number of subjects, and age of the tDCS and sham groups; (III) information on the intervention, including the type of stimulation (anode/cathode), blinding, offline and online intensity and current level, as well as the methods used to carry out the tDCS intervention.

### 2.5 Quality assessment

To assess the quality of the included studies, a 7-point scale was used with a score range of 0 to 7 points. This scale included seven items: (I) sequence generation (randomization and counterbalancing); (II) random assignment of groups; (III) allocation concealment or blinded outcome assessment; (IV) blinding of participants; (V) calculation of sample size; (VI) adherence to human ethics, protection laws, and regulations; and (VII) declaration of relevant conflicts of interest. Each item was given a quality score, and the greater the score, the higher the study quality.

### 2.6 Statistical analysis

The global effect of tDCS on emotion was assessed using a random-effects model to account for heterogeneity. This analysis utilized the standardized mean difference (SMD) and 95% confidence intervals (CIs) as specified in the Cochrane Handbook for Systematic Reviews of Interventions. The I^2^ statistic was employed to assess heterogeneity, with low, moderate, and considerable heterogeneity defined as values of 25%, 50%, and 75%, respectively. The I² value indicates the extent of heterogeneity among the included studies, beyond chance variability (Higgins and Thompson 2002). We compared the emotional processing between tDCS and sham groups for the experiments.

## 3. Results

### 3.1 Study inclusion

Initially, 3355 articles were found using the terms in Scopus, WOS, and PubMed databases. The number of articles was reduced to 1833 after duplicates were eliminated. Based on the previously established inclusion and exclusion criteria, 1522 articles were excluded due to various factors, such as being a review, conference, or case report study, not involving human subjects, or not related to emotion as assessed by the title and abstract. For subsequent selection, the 133 remaining studies were downloaded, and 101 additional studies were excluded based on inclusion (or exclusion) criteria described previously or because they did not report data to create summary statistics needed for the meta-analysis after reading their full text. Eventually, 32 studies were included in this systematic review and meta-analysis study (Fig 1).

**Fig. 1.**
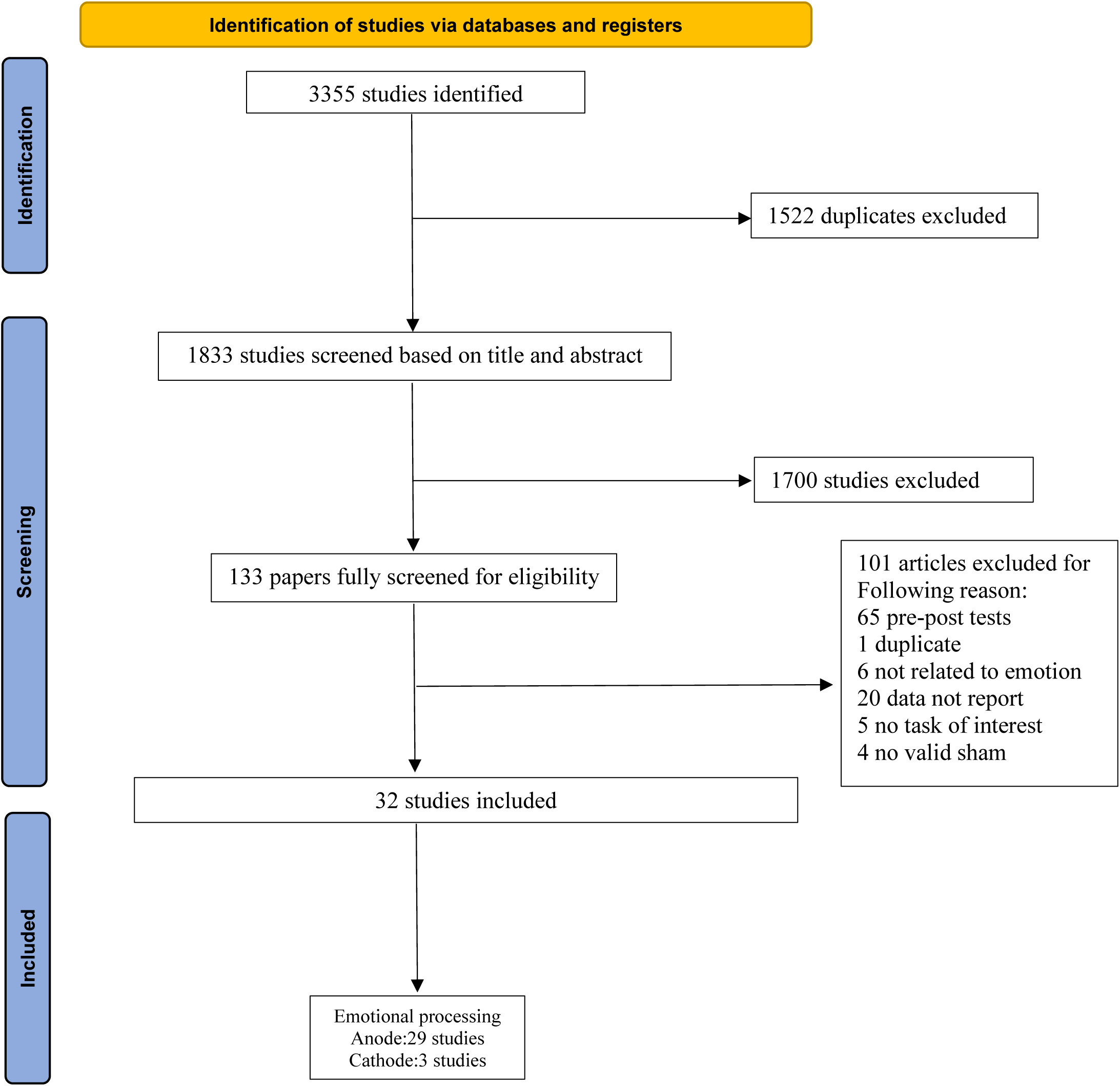
Flow chart of the meta-analysis procedure based on the PRISMA guideline

### 3.2 Study characteristics

There were 2107 participants in 32 articles, of which 1056 participants were in the sham group also, and 1051 subjects were in the tDCS group (Table 1–2). All 32 articles included in the review provided the age range of their participants, which ranged from 6 to 84 years old. From the 32 studies, the stimulation duration was 5 to 35 minutes, the current level was 0.5 to 2.5 mA. Thirteen studies were double-blind, nine studies were single-blind, and the blinding status of ten studies was not reported. Sham interventions in the studies consisted of a fictitious stimulus.

**Table 1.** Characteristics of the included studies based on the effect of anodal tDCS on Emotional processing.

| Study | Subjects | Age (y) | tDCS protocol |  |  |  |  |  |  |  | Test |
| --- | --- | --- | --- | --- | --- | --- | --- | --- | --- | --- | --- |
|  |  |  | Anode | Cathode | Size (cm) | Current (mA) | Session | Blinding | Timing | During (min) |  |
| (Balzarotti and Colombo 2016) | Healthy | 24.57 | F3 | Ipsilateral mastoid | 5x5 | 1.5. | NR | NR | Online | 20 | Filler |
| (Brennan, McLoughlin et al. 2017) | Healthy | 19 - 58 | F3 | Contralateral supraorbital | 5x7 | 1.5 | NR | SB | Online | 30 | Tapping |
| (Colombo, Balzarotti et al. 2016) | Healthy | 21.-55 | F3 | Ipsilateral mastoid | 5x5 | 1.5 | NR | SB | Online | 15 | Filler |
| (Conson, Errico et al. 2015) | Healthy | 22-30 | F3 | F4 | 5x7 | 1 | NR | NR | Online | 15 | Visual Perspective Taking |
| (Dambacher, Schuhmann et al. 2015) | Healthy | 22.14 | F4 | Left eyebrow | 5x7 | 2 | NR | NR | Online | 20 | Taylor Aggression Paradigm Reactive |
| (Faehling and Plewnia 2016) | Healthy | 84.04 | F3 | LDLPFC | 5x7 | 1.5 | NR | SB | Online | 28 | Working Memory |
| (Nejati, Sarraj Khorrami et al. 2021) | Healthy | 6 -12 | Fp2 | F3 | 4 x 6 | 1 | 3 | SB | Online | 15 | Chocolate delay discounting |
| (Nejati, Majdi et al. 2021) | Healthy | 20-30 | RDLPFC | LDLPFC | 5x5 | 1.5 | 3 | SB | online | 15 | Emotional picture rating |
| (Nord, Forster et al. 2017) | Healthy | 25.6 | LDLPFC | Ipsilateral shoulder | 5 x 7 | 1 | 2 | DB | Online | 20 | Distractibility task |
| (Qiao, Hu et al. 2020) | Autistic | 18- 22 | Cp6 | C4, p4, T8, p8 | 5 x 7 | 2 | NR | SB | Online | 20 | Eye-tracking apparatus |
| (Schroeder, Ehli et al. 2015) | Healthy | 31.3 | LDLPFC | Arm | 5 x 7 | 1 | 1 | DB | Online | 20 | Electrodermal Recordings |
| (Trémolière, Maheux-Caron et al. 2018) | Healthy | 23.71 | LDLPFC | Supraorbital | 5 x 7 | 2 | 1 | NR | Online | 20 | Reasoning task |
| (Wang, Jung et al. 2020) | Healthy | 20.31 | F3, F4 | F4, F3 | NR | 2 | NR | NR | Online | 20 | Trust Game |
| (Ayache, Palm et al. 2016) | MS Patients | 18-70 | F3 | Over the right supraorbital | 5 x 5 | 2 | 3 | NR | Offline | 20 | Attention Network |
| (Boggio, Ferrucci et al. 2006) | Healthy | 18 - 35 | C3 | Supraorbital opposite side. | 5x7 | 2 | 1 | DB | Online | 5 | Human pain skill |
| (Grant, Geoghegan et al. 2020) | Healthy | 18 - 50 | LDLPFC | Left superior trapezius | 4 x 6 cm | 2 | 1 | NR | Offline | 20 | Emotion regulation |
| (Donaldson, Lee et al. 2019) | Healthy | 18-40 | P6 | NR | NR | 2 | 1 | DB | Offline | 20 | Fear Surprise |
| (Feesser, Fan et al. 2015) | Psychiatric Disorders | 20-47 | RDLPFC | Left supraorbital region | 5 x 7 | 1.5 | 2 | DB | Offline | 20 | Skin conductance responses |
| (Gilam, Abend et al. 2018) | Healthy | 16±3.63 | Over the forehead | Right shoulder | 5 x 7 | 1.5 | 2 | DB | Offline | 22 | Anger-infused Ultimatum Game |
| (Herrmann, Adelman et al. 2016) | Healthy | 18-35 | F8 | Fp1 | 5 x 7 | 2 | NR | DB | Offline | 20 | Skin conductance responses |
| (Jafari, Pormohammad et al. 2021) | Social Anxiety Disorder | 18-50 | LDLPFC | Fpz | 5 x 7 | 2<br>1 | 10 | DB | Offline | 20 | Attentional bias |
| (Koenigs, Huey et al. 2008) | Healthy | NR | Forehead | Arm | 5 x 5 | 2.5 | 3 | DB | Offline | 35 | Skin conductance responses |
| (Kuehne, Weiskittel et al. 2019) | Healthy | 24.4 | LDLPFC | NR | NR | 0.5 | 2 | NR | Offline | 20 | Emotional face-word stroop |
| (Maeoka, Matsuo et al. 2012) | Healthy | 22-24 | F3 | Over the supraorbital | 5 x 7 | 1 | 2 | SB | Offline | 20 | Oddball |
| (Sagliano, Atripaldi et al. 2019) | Healthy | 18-31 | LDLPFC | RDLPCF | 5 x 5 | 1 | 3 | NR | Offline | 15 | Classic Posner |
| (Sanchez, Vanderhasselt et al. 2016) | Healthy | 23.27 | DLPFC | Contra lateral supraorbital | 5 x 7 | 2 | 2 | SB | Offline | 20 | Attention |
| (Sánchez-López, Hernández et al. 2016) | Healthy | 23.4 | LDLPFC | RDLPCF | 5 x 7 | 2 | 2 | SB | Offline | 20 | Attentional engagement-disengagement |
| (Marie, Dominguez-Vega et al. 2016) | Depressed participants | 18-65 | LDLPFC | Right Supraorbital | 5x5 | 2 | SB | DB | Online | 15 | Go/no-go |
| (Weigand, Thomas et al. 2021) | Healthy | 19-30 | rSMG | Contralateral supraorbital | 5 x 7 | 1 | 2 | DB | Offline | 20 | SOFE |

**Table 2.** Characteristics of the included studies based on the effect of cathodal tDCS on Emotional processing.

| Study | Subjects | Age (y) | tDCS protocol |  |  |  |  |  |  |  | Test |
| --- | --- | --- | --- | --- | --- | --- | --- | --- | --- | --- | --- |
|  |  |  | Anode | Cathode | Size (cm) | Current<sup>(mA)</sup> | Session | Blinding | Timing | During<sup>(min)</sup> |  |
| (Sagliano, Atripaldi et al. 2019) | Healthy | 18-31 | LDLPFC | RDLPFC | 5 x 5 | 1 | 3 | NR | Offline | 15 | Classic Posner |
| (Koenigs, Huey et al. 2008) | Healthy | NR | Forehead | Arm | 5 x 5 | 2.5 | 3 | DB | Offline | 35 | Skin conductance responses |
| (Donaldson, Lee et al. 2019) | Healthy | 18-40 | P6 | NR | NR | 2 | 1 | DB | Offline | 20 | Fear Surprise |
| (Johnson and Durrant 2018) | Healthy | 18-22 | NR | NR | NR | NR | NR | NR | offline | NR | Serial reaction time |
| (Peña-Gómez, Vidal-Pineiro et al. 2011) | Healthy | 22.93 | F3 | C4 | 5 x 7 | 1 | 2 | NR | Offline | 20 | Emotional valence rating |
| (Volkenstein, Zeiller et al. 2014) | Major Depression | 19-54 | LDLPFC | Contralateral deltoid muscle | 5 x 7 | 1 | 2 | DB | Offline | 20 | Delayed response working memory |

### 3.3 Articles quality and publication bias

The study quality score ranged from one to seven out of a maximum of 7 points. Two articles received 1 point; three articles received 2; four articles received 3; eight articles received 4; eight articles received 5; five articles received 6; and two articles received 7. Sequence generation or counterbalancing had been reported in 10 articles, and random assignment of groups had been reported in 24 studies. In 23 studies, allocation concealment or outcome assessment had been mentioned. Blinding participants had been reported in 23 studies. Calculation of sample size had been reported in 6 studies. Conformity to human ethics has been reported in 28 studies. Declaration of any relevant conflicts of interest had been reported in 6 studies (Table 3).

**Table 3.** Articles quality and publication bias.

| Study | (A) | (B) | (C) | (D) | (E) | (F) | (G) | Total |
| --- | --- | --- | --- | --- | --- | --- | --- | --- |
| (Ayache, Palm et al. 2016) |  | . | . | . |  | . | . | 5 |
| (Boggio, Ferrucci et al. 2006) | . | . | . | . |  | . |  | 5 |
| (Grant, Geoghegan et al. 2020) | . | . |  |  |  | . | . | 4 |
| (Donaldson, Lee et al. 2019) | . | . | . | . |  | . |  | 5 |
| (Feesser, Fan et al. 2015) | . | . | . | . |  | . | . | 6 |
| (Gilam, Abend et al. 2018) |  | . | . | . |  | . | . | 5 |
| (Herrmann, Adelman et al. 2016) | . | . | . | . |  | . | . | 6 |
| (Jafari, Pormohammad et al. 2021) | . | . | . | . | . | . | . | 7 |
| (Johnson and Durrant 2018) |  |  |  |  |  | . | . | 2 |
| (Koenigs, Huey et al. 2008) |  |  | . | . |  |  |  | 2 |
| (Kuehne, Weiskittel et al. 2019) |  |  |  |  |  | . | . | 2 |
| (Maeoka, Matsuo et al. 2012) |  | . | . | . |  |  |  | 3 |
| (Peña-Gómez, Vidal-Pineiro et al. 2011) |  | . |  |  |  | . | . | 3 |
| (Sagliano, Atripaldi et al. 2019) |  |  |  |  |  | . |  | 1 |
| (Sanchez, Vanderhasselt et al. 2016) |  | . | . | . |  | . |  | 4 |
| (Sánchez-López, Hernández et al. 2016) |  | . | . | . |  | . |  | 4 |
| (Marie, Dominguez-Vega et al. 2016) |  |  | . | . |  | . |  | 3 |
| (Weigand, Thomas et al. 2021) | . | . | . | . | . | . | . | 7 |
| (Wolkenstein, Zeiller et al. 2014) |  | . | . | . |  | . | . | 5 |
| (Balzarotti and Colombo 2016) |  | . | . | . |  | . | . | 5 |
| (Brennan, McLoughlin et al. 2017) |  | . | . | . |  | . | . | 5 |
| (Colombo, Balzarotti et al. 2016) |  | . |  |  |  |  |  | 1 |
| (Conson, Errico et al. 2015) | . | . |  |  |  | . | . | 4 |
| (Dambacher, Schuhmann et al. 2015) | . | . |  |  |  | . | . | 4 |
| (Faehling and Plewnia 2016) |  | . | . | . |  |  | . | 4 |
| (Nejati, Majdi et al. 2021) |  | . | . | . | . | . | . | 6 |
| (Nejati, Sarraj Khorrami et al. 2021) |  | . | . | . | . | . | . | 6 |
| (Nord, Forster et al. 2017) |  | . | . | . |  | . | . | 4 |
| (Qiao, Hu et al. 2020) | . |  | . | . |  | . | . | 5 |
| (Schroeder et al., 2015) |  |  | . | . |  | . | . | 4 |
| (Trémolière, Maheux-Caron et al. 2018) |  |  |  |  | . | . | . | 3 |
| (Wang, Jung et al. 2020) |  | . | . | . | . | . | . | 6 |
| Sum | 10 | 24 | 23 | 23 | 6 | 28 | 23 |  |

(A) sequence generation (randomization and counterbalancing); (B) random assignment of groups; (C) allocation concealment or blinded outcome assessment; (D) blinding of participants; (E) calculation of sample size; (F) adherence to human ethics, protection laws, and regulations; and (G) declaration of relevant conflicts of interest.

### 3.4 Assessment of bias

To address publication bias funnel plots of reported effect sizes were examined. Funnel plots show the effect size variability in each study as a function of its mean. In the absence of bias, the data points are expected to be roughly symmetric around the mean effect size across all studies (Tang & Liu, 2000). Fig 2–3 shows that the tDCS effect on emotional processing was relatively symmetric around the mean.

**Fig. 2.**
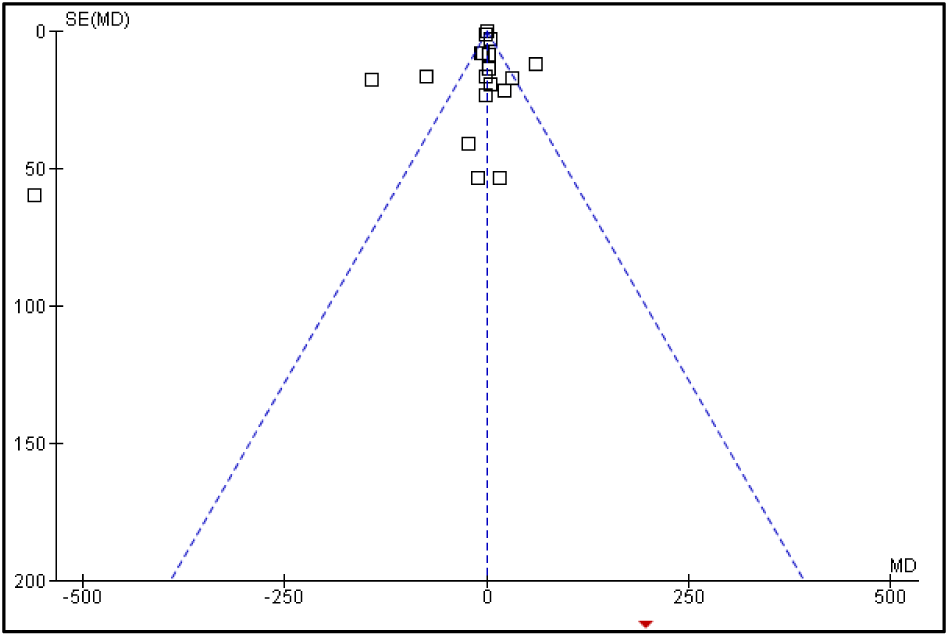
Funnel plot depicting anodal tDCS effects on reaction time. The X-axis represents standardized mean difference (SMD), while the Y-axis represents standard error (SE) of SMD.

**Fig. 3.**
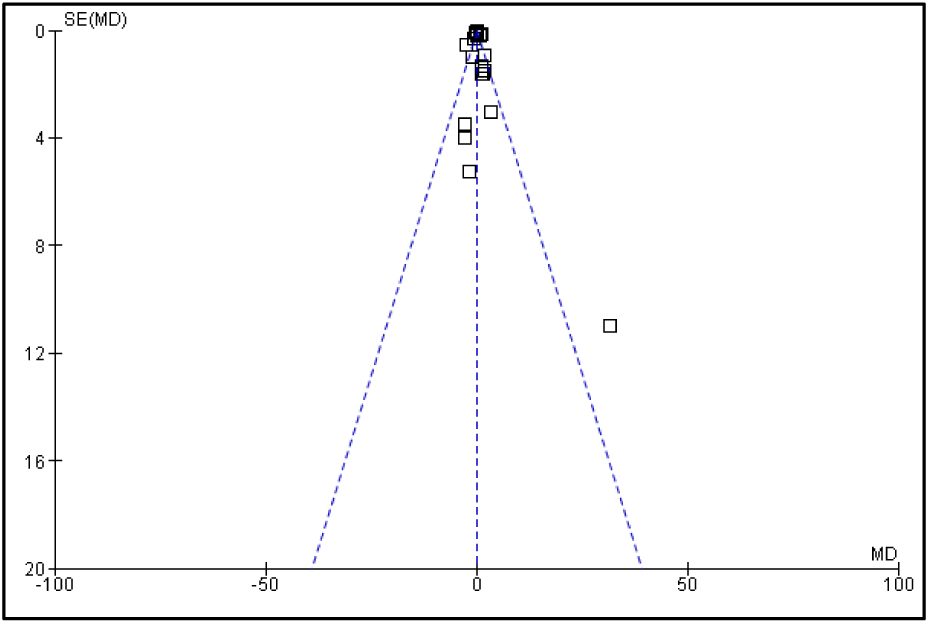
Funnel plot depicting anodal tDCS effects on accuracy. The X-axis represents standardized mean difference (SMD), while the Y-axis represents standard error (SE) of SMD

### 3.5 tDCS Effect on Emotional Processing

#### Effect of anodal tDCS: RT

The effect of anodal tDCS on emotion RT is reported in twenty-two studies (Table 1). Results show a significant effect of anodal tDCS on RT as compared to the sham group (n = 1166, SMD= −0.10, 95 % CI: −0.41 ~ 0.22, P < 0.00001; heterogeneity χ2 = 137.09, I2 = 85 %, Fig 4A and S1).

**Fig. 4.**
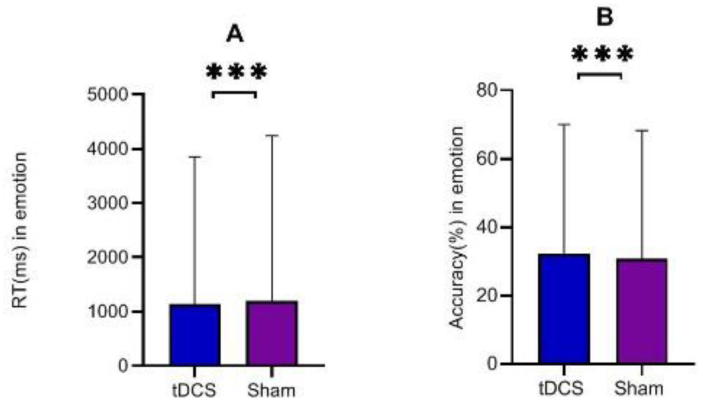
The meta-analysis of the anodal tDCS effect on A) RT (ms), and B) accuracy in emotion. Error bars are SEM. The symbols *, **, and *** indicate statistical significance levels at p < 0.05, p < 0.01, and p < 0.001, respectively.

#### Effect of anodal tDCS: Accuracy

In twenty-one studies (Table 1), anodal tDCS was applied to improve the accuracy of emotional processing, with a significant impact compared to the sham group (n = 941, SMD= 0.11, 95 % CI: −0.22 ~ 0.43, P < 0.00001; heterogeneity χ2 = 115.17, I2 = 83 %, Fig 4B and S2).

#### Effect of cathodal tDCS: RT

Five studies applied cathodal tDCS to modulate RT for emotional processing. The use of RT as a measure revealed a significant effect of cathodal tDCS on emotion RT compared to the sham group (n = 204, SMD= −0.46, 95 % CI: −1.44 ~ 0.53, P < 0.00001; heterogeneity χ2 = 43.28, I2 = 91 %, Fig.5A and S3).

**Fig. 5.**
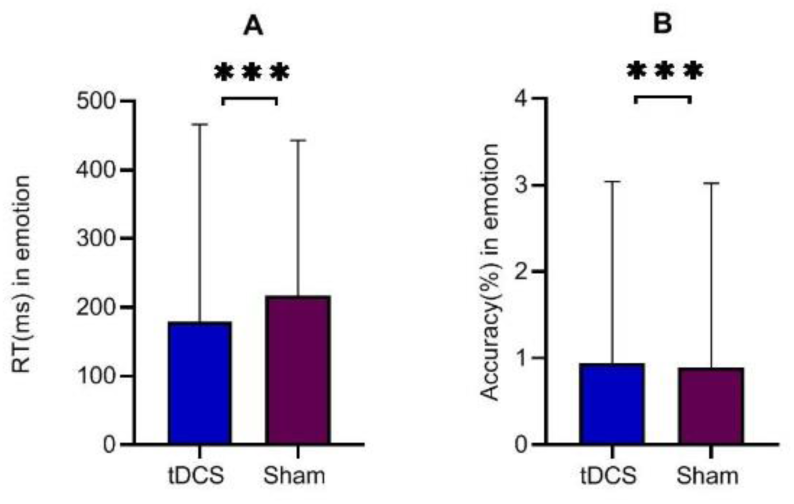
The meta-analysis of the cathodal tDCS effect on A) RT (ms), and B) accuracy in emotion. Error bars are SEM. The symbols *, **, and *** indicate statistical significance levels at p < 0.05, p < 0.01, and p < 0.001, respectively.

#### Effect of cathodal tDCS: Accuracy

Six studies utilized cathodal tDCS to enhance the accuracy of emotional processing, with a significant effect observed in comparison to the sham group (n = 246, SMD= 0.61, 95 % CI: −0.10 ~ 1.31, P < 0.00001; heterogeneity χ2 = 34.78, I2 = 86 %, Fig.5B and S4).

### 3.6 The Influence of Stimulation Parameters of tDCS on Emotional Processing

#### 3.6.1 Stimulation Sites

The study results revealed significant differences in how stimulation parameters affected emotion. Frontal tDCS had a significantly greater impact on RT (n = 1296, SMD= −0.07, 95 % CI: −0.34 ~ 0.20, P < 0.00001; heterogeneity χ2 = 139.85, I2 = 81 %, Fig. 6A and S5) and accuracy (n = 859, SMD= 0.17, 95 % CI: −0.17 ~ 0.52, P < 0.0001; heterogeneity χ2 = 106.12, I2 = 83 %, Fig. 6B and S6) compared to tDCS applied to other regions.

**Fig. 6.**
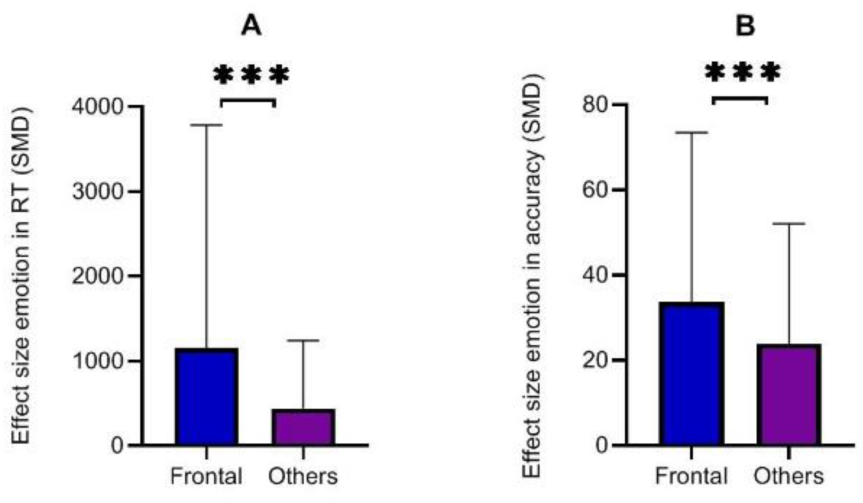
The subgroup meta-analysis based on the influence of stimulation site of tDCS effect on A) RT (ms), and B) accuracy in emotion. Error bars are SEM. The symbols *, **, and *** indicate statistical significance levels at p < 0.05, p < 0.01, and p < 0.001, respectively.

#### 3.6.2 Stimulation Duration

Subgroup analysis of stimulation time revealed a significant difference in RT (n = 1180, SMD= −0.12, 95 % CI: −0.43~ 0.20, P < 0.00001; heterogeneity χ2 = 136.15, I2 = 85 %, Fig. 7A and S7) and accuracy (n = 941, SMD= 0.11, 95 % CI: −0.22 ~ 0.43, P < 0.00001; heterogeneity χ2 = 115.17, I2 = 83 %, Fig. 7B and S8) between stimulation durations of 15 minutes, 20 minutes, and more than 20 minutes.

**Fig. 7.**
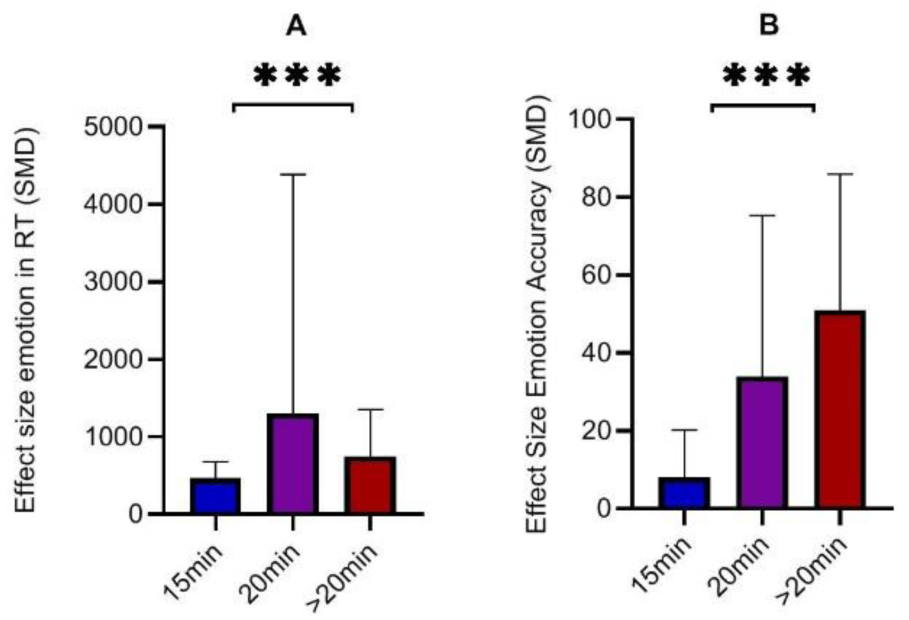
The subgroup meta-analysis based on the influence of during stimulation of tDCS effect on A) RT (ms), and B) accuracy in emotion. Error bars are SEM. The symbols *, **, and *** indicate statistical significance levels at p < 0.05, p < 0.01, and p < 0.001, respectively.

#### 3.6.3 Electrical Current Intensities

Subgroup analysis of tDCS current intensity revealed significant differences in RT (n = 865, SMD= −0.10, 95 % CI: −0.40 ~ 0.21, P < 0.00001; heterogeneity χ2 = 137.32, I2 = 84 %, Fig. 8A and S9) and accuracy (n = 859, SMD= 0.18, 95 % CI: −0.16 ~ 0.52, P < 0.00001; heterogeneity χ2 = 105.87, I2 = 83 %, Fig. 8B and S10) between current intensities of 1 mA, 1.5 mA, and 2 mA. Notably, tDCS at 2 mA had a greater impact on emotional processing compared to the lower current intensities.

**Fig. 8.**
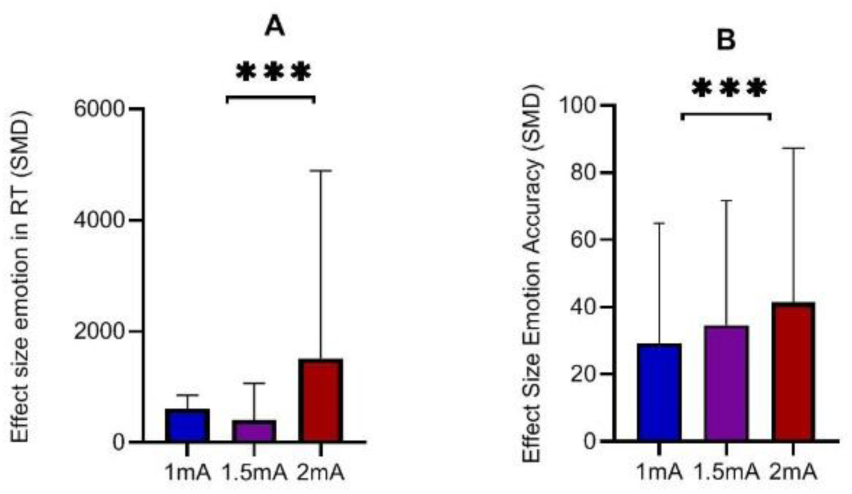
The subgroup meta-analysis based on the influence of the current level of tDCS effect on A) RT (ms), and B) accuracy in emotion. Error bars are SEM. The symbols *, **, and *** indicate statistical significance levels at p < 0.05, p < 0.01, and p < 0.001, respectively.

#### 3.6.4 Participant Type: Clinical vs. Healthy Studies

The analysis of population-based tDCS studies (clinical vs. healthy) showed a significant effect on RT (n = 1066, SMD= −0.11, 95 % CI: 0.42 ~ 0.21, P < 0.00001; heterogeneity χ2 = 135.03, I2 = 83 %, Fig. 9A and S11) and accuracy (n = 863, SMD= 0.17, 95 % CI: −0.17 ~ 0.51, P < 0.00001; heterogeneity χ2 = 106.28, I2 = 83 %, Fig. 9B and S12) in studies with healthy participants compared to studies with clinical participants.

**Fig. 9.**
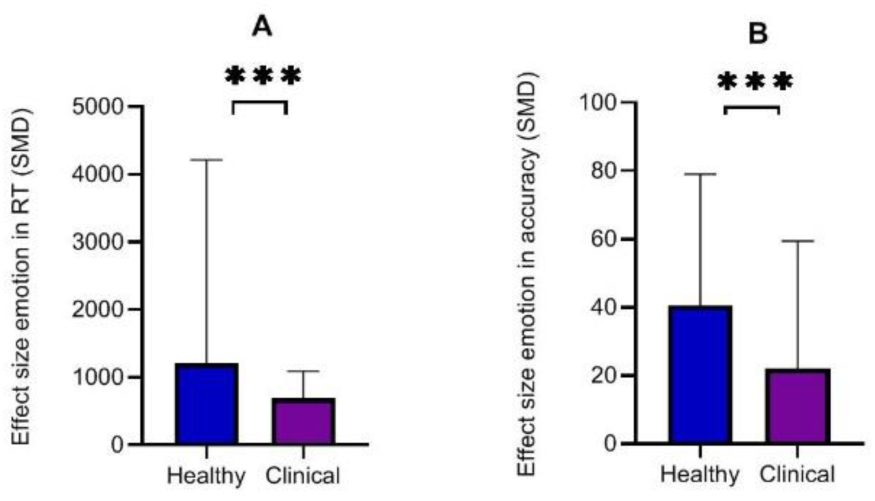
The subgroup meta-analysis based on the influence of population of tDCS effect on A) RT (ms), and B) accuracy in emotion. Error bars are SEM. The symbols *, **, and *** indicate statistical significance levels at p < 0.05, p < 0.01, and p < 0.001, respectively.

#### 3.6.5 Timing of Stimulation: Online vs. Offline Studies

The subgroup analysis focusing on the timing of tDCS (online vs. offline) showed a significant effect on RT (n = 1256, SMD= −0.08, 95 % CI: −0.37 ~ 0.21, P < 0.00001; heterogeneity χ2 = 138.59, I2 = 83 %, Fig. 10A and S13) and accuracy (n = 941, SMD= 0.11, 95 % CI: −0.22 ~ 0.43, P < 0.00001; heterogeneity χ2 = 115.17, I2 = 83 %, Fig. 10B and S14) in online studies compared to offline studies.

**Fig. 10.**
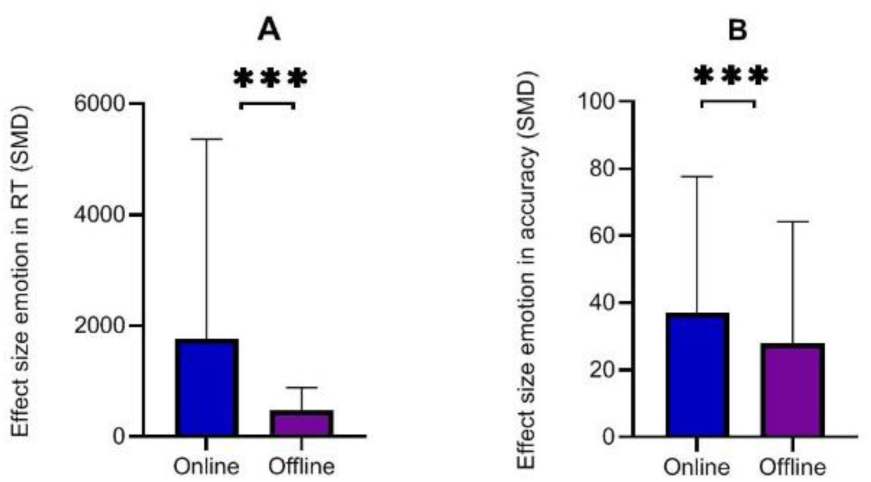
The subgroup meta-analysis based on the influence of timing of tDCS on RT (SMD) in A) Emotion and accuracy (SME) in B motion. Error bars are SEM. The symbols *, **, and *** indicate statistical significance levels at p < 0.05, p < 0.01, and p < 0.001, respectively

#### 3.6.6 Age Group: Young vs. Older

The findings regarding the influence of tDCS parameters, such as age, on emotional processing revealed significant differences. The analysis revealed a significant difference in RT (n = 1158, SMD= −0.13, 95% CI: −0.44, ~0.19, P < 0.00001; heterogeneity χ2 = 133.90, I2 = 84 %, Fig. 11A and S15) and accuracy (n = 891, SMD= 0.16, 95% CI: ~-0.17, ~0.48, P < 0.00001; heterogeneity χ2 = 106.31, I2 = 82%, Fig. 11B and S16) between ages of <25 years as compared to the > 25-year-old group.

**Fig. 11.**
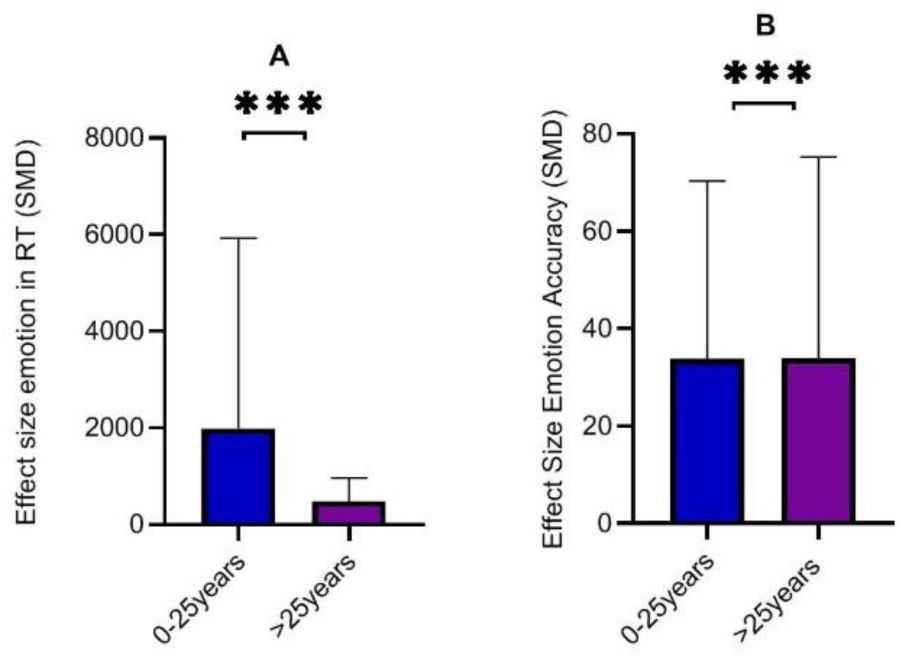
The subgroup meta-analysis based on the influence of age of tDCS effect on A) RT (ms), and B) accuracy in emotion. Error bars are SEM. The symbols *, **, and *** indicate statistical significance levels at p < 0.05, p < 0.01, and p < 0.001, respectively.

## 4. Discussion

Previous research on the effects of tDCS on emotional processing has yielded inconsistent findings, largely due to variations in study quality, methodology, and stimulation parameters. To address these inconsistencies and assess the robustness of tDCS effects, we conducted a meta-analysis of 32 studies that investigated the impact of anodal and cathodal tDCS on emotional processing. These studies varied across multiple dimensions, including stimulation site, current intensity, duration, total stimulation time, type of stimulation (online vs. offline), outcome measures, and participant characteristics. Our study illustrates the significant effects of both anodal and cathodal tDCS on emotion RT and accuracy (Fig 4 and Fig 5). Additionally, the results highlight significant differences in how tDCS stimulation parameters influence emotional processing, demonstrating that frontal tDCS enhances emotional outcomes more effectively than stimulation of other brain regions (Fig 6). Key findings include that longer stimulation durations (over 20 minutes, Fig 7) and higher current intensities (2 mA) yield better results (Fig 8), with online tDCS being more effective than offline stimulation (Fig 10), particularly in participants under 25 and in non-clinical populations (Fig 9). Our results reveal that tDCS can effectively enhance emotional processing, offering valuable insights into the potential benefits of this method for emotion improvement.

Our findings demonstrate that anodal tDCS significantly improves both RT and accuracy in emotional processing, aligning with previous studies (Dambacher, Schuhmann et al. 2015, Nejati, Sarraj Khorrami et al. 2021, Qiu, He et al. 2023). Interestingly, we also observed significant effects of cathodal tDCS. Anodal tDCS is thought to enhance neural excitability in targeted brain regions by depolarizing neurons (Fig 12), making them more likely to fire and thereby potentially improving cognitive task performance. This increase in excitability may streamline neural networks involved in emotional processing, as seen in the faster RT and improved accuracy observed in studies using anodal tDCS. Conversely, cathodal tDCS, which hyperpolarizes neurons and reduces excitability, has also been found to positively influence emotional processing, likely by fine-tuning neural networks and enhancing task precision (Nitsche, Kuo et al. 2015). Many studies associate tDCS-induced changes in neural activity with neuroplasticity, though they also highlight the concept of brain flexibility—whereby the brain maximizes performance based on current conditions—as a possible mechanism for these enhancements. Recent research points to complex molecular and cellular processes underlying both short- and long-term synaptic changes with non-invasive brain stimulation, including gene regulation, protein modulation, synaptic transmission, neurotrophic factor adjustments, and glial function modulation. These mechanisms collectively suggest that tDCS impacts emotional processing by boosting activity in the prefrontal cortex (PFC) (Shaker, Sawan et al. 2018).

**Fig. 12.**
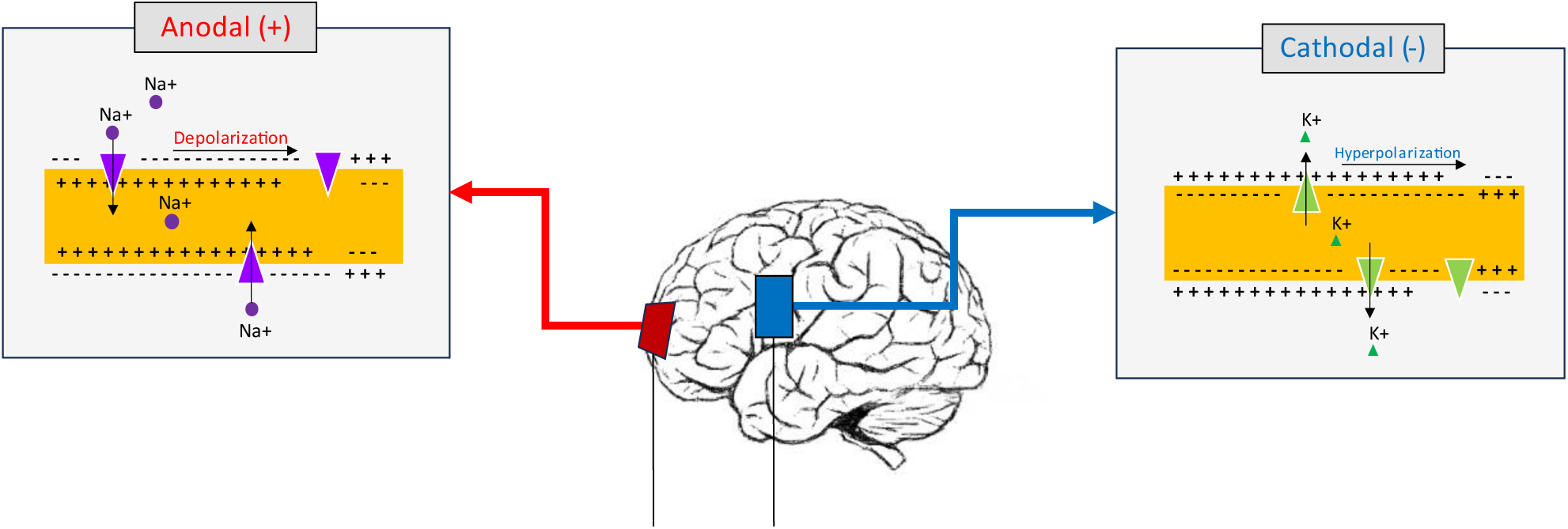
Mechanisms of action induced by tDCS. The red anodal electrode delivers a mild electrical current toward the blue cathodal electrode. On the right side, the cathodal electrode induces hyperpolarization in the neurons, leading to a decrease in neuronal activity. On the left side, the anodal electrode causes the opposite effect, depolarizing the neurons and increasing their activity.

The current study thoroughly explores the intricate relationships among different stimulation parameters used in tDCS and their complex effects on emotional processing. Firstly, the significant impact of frontal tDCS on emotional processing underscores the prefrontal cortex’s pivotal role in emotion regulation (Estaji, Hosseinzadeh et al. 2024), suggesting that targeting this region could enhance therapeutic outcomes for emotional disorders. The analysis of stimulation duration indicated that longer durations (more than 20 minutes) were more effective (Dos Reis, Pereira Generoso et al. 2024, Zheng, Wong et al. 2024), suggesting a dose-response relationship where extended stimulation may facilitate more profound neural modulation. Additionally, higher current intensities, particularly 2 mA, were found to have a greater impact on emotional processing (Nitsche, Koschack et al. 2012, Allaert, Sanchez-Lopez et al. 2019), highlighting the importance of adjusting current intensity to achieve desired effects. The differential effects observed between clinical and healthy participants suggest that tDCS may be more effective in modulating emotional processing in non-clinical populations (Zheng, Wong et al. 2024), possibly due to baseline differences in neural plasticity (Sierawska, Splittgerber et al. 2023) or emotional regulation mechanisms. Furthermore, the greater efficacy of online tDCS compared to offline tDCS on emotional processing (Qiu, He et al. 2023), suggests that concurrent cognitive engagement may enhance the neuromodulatory effects of tDCS. Lastly, age-related differences, with younger participants showing more significant changes, indicate that age-related neural plasticity may influence tDCS outcomes (Greeley, Barnhoorn et al. 2022), necessitating age-specific adjustments in stimulation protocols. These findings collectively emphasize the need for personalized tDCS parameters to maximize therapeutic benefits in emotional processing.

Our study also emphasizes the need for careful consideration of study quality in meta-analyses. While we did not exclude studies based on quality scores, we recognize that study quality can influence the overall findings. To mitigate this potential limitation, future research should incorporate sensitivity analyses to assess the impact of lower-quality studies on the meta-analytic outcomes. This approach would allow for a more nuanced understanding of the effects of study quality and enhance the robustness of the conclusions drawn from meta-analyses.

In summary, our meta-analysis underscores the significant impact of tDCS parameters on emotional processing, revealing crucial insights into how stimulation site, duration, current intensity, age, and participant characteristics influence outcomes. These findings advocate for a tailored approach in tDCS applications to optimize emotional processing enhancements and suggest further investigation into the underlying mechanisms involved.

## Author Contribution

Conceived and designed the experiments: FA and AN. Performed the experiments: FA and AN. Analyzed the data: FA and AN. Contributed reagents/materials/analysis tools: FA and AN. Wrote the paper: FA and AN. Agreed with the manuscript’s results and conclusions: FA and AN. All authors read and approved the final manuscript.

## Availability of Data and Materials

The datasets supporting the conclusions of this article are included in the supplementary file.

## Declarations Ethical Approval

Not Applicable.

## Data Availability

The datasets supporting the conclusions of this article are included in the supplementary file.

