## Supplementary material for "The Efficacy of transcranial Direct Current Stimulation (tDCS) on Emotional Processing": supplimentry

| Study or Subgroup | tDCS Mean | tDCS SD | tDCS Total | Sham Mean | Sham SD | Sham Total | Weight | Std. Mean Difference IV, Random, 95% CI |
|---|---|---|---|---|---|---|---|---|
| Donaldson et al, 2019off | -618.944 | 236.218 | 17 | -57.727 | 72.658 | 18 | 3.5% | -3.18 [-4.21, -2.15] |
| Jafari et al,2021 2maoff | 427.16 | 46.45 | 13 | 569.58 | 45.11 | 14 | 3.2% | -3.02 [-4.17, -1.87] |
| Jafari et al,2021off | 495.08 | 42.27 | 14 | 569.58 | 45.11 | 14 | 4.0% | -1.65 [-2.53, -0.78] |
| Colombo et al,2016 | 4.18 | 3.1307 | 10 | 6.05 | 3.1307 | 10 | 3.9% | -0.57 [-1.47, 0.33] |
| Tremoliere et al,2018 | 13,000 | 6,392 | 34 | 14,693 | 9,599 | 34 | 5.0% | -0.21 [-0.68, 0.27] |
| Ayache et al, 2016off | 742.3 | 99.7 | 16 | 766.1 | 130.3 | 16 | 4.5% | -0.20 [-0.89, 0.49] |
| Clarke et al,2020 off | 972.21 | 50.33 | 79 | 979 | 50.13 | 79 | 5.4% | -0.13 [-0.45, 0.18] |
| Dambacher et al,2015 | 3.84 | 1.16 | 16 | 4 | 1.33 | 16 | 4.5% | -0.12 [-0.82, 0.57] |
| Clarke et al,2020 benign | 949.68 | 52.14 | 79 | 955.82 | 52.04 | 79 | 5.4% | -0.12 [-0.43, 0.19] |
| Conson et al,2015 | 616.69 | 150.4 | 16 | 627.82 | 151.56 | 16 | 4.5% | -0.07 [-0.77, 0.62] |
| Sanchez Lopez et al,2016R | 340 | 54 | 27 | 342 | 69 | 27 | 4.9% | -0.03 [-0.57, 0.50] |
| Sanchezlopez etal,2018of | 390 | 94 | 27 | 391 | 79 | 27 | 4.9% | -0.01 [-0.54, 0.52] |
| Nord et al,2017 | 479.59 | 56.0674 | 31 | 477.19 | 53.3949 | 31 | 5.0% | 0.04 [-0.45, 0.54] |
| Boggio et al,2006 off | 209.15 | 77.62 | 26 | 205.45 | 61.62 | 26 | 4.9% | 0.05 [-0.49, 0.60] |
| Sagliano et al,2017off | 319.13 | 27.45 | 20 | 317.12 | 28.97 | 20 | 4.7% | 0.07 [-0.55, 0.69] |
| Marie Anne et al,2016off | 784.79 | 225.1 | 35 | 768.59 | 222.39 | 35 | 5.1% | 0.07 [-0.40, 0.54] |
| Qiao et al,2020 | 1,269.45 | 942.31 | 14 | 1,073.51 | 657.13 | 15 | 4.4% | 0.24 [-0.50, 0.97] |
| Kuehne et al,2019off | 723.05 | 72.29 | 18 | 700.9 | 58.84 | 18 | 4.6% | 0.33 [-0.33, 0.99] |
| Brennan et al,2016 | 63.8 | 10.24 | 20 | 60.35 | 8.7 | 20 | 4.7% | 0.36 [-0.27, 0.98] |
| Sanchez et al,2016off | 379.16 | 71.72 | 30 | 348.92 | 63.26 | 30 | 4.9% | 0.44 [-0.07, 0.95] |
| Faehling and Plewnig,2016 | 1,170.09 | 45.57 | 21 | 1,110.98 | 34.62 | 22 | 4.5% | 1.44 [0.76, 2.12] |
| Fan et al,2020 | 2,374.777 | 437.793 | 18 | 1,448.777 | 124.477 | 18 | 3.8% | 2.81 [1.86, 3.76] |
| **Total (95% CI)** | | | **581** | | | **585** | **100.0%** | **-0.10 [-0.41, 0.22]** |

Heterogeneity: Tau² = 0.46; Chi² = 137.09, df = 21 (P < 0.00001); I² = 85%
Test for overall effect: Z = 0.59 (P = 0.55)

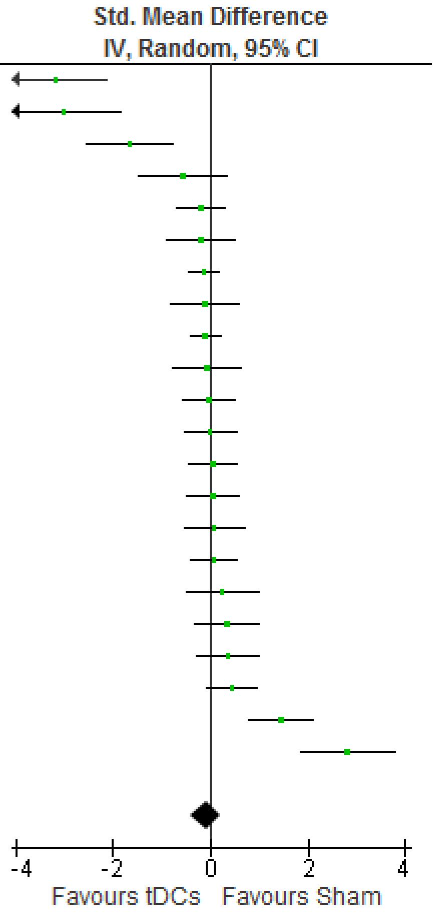


**Fig. S1** The meta-analysis of the anodal tDCS effect on emotion RT.

| Study or Subgroup | tDCS Mean | tDCS SD | tDCS Total | Sham Mean | Sham SD | Sham Total | Weight | Std. Mean Difference IV, Random, 95% CI |
|---|---|---|---|---|---|---|---|---|
| Faehling and Plewnig,2016 | 79.29 | 1.72 | 21 | 81.7 | 1.74 | 22 | 4.7% | -1.37 [-2.04, -0.70] |
| Feeser et al, 2013off | 0.26 | 0.27 | 48 | 0.56 | 0.36 | 48 | 5.4% | -0.94 [-1.36, -0.51] |
| Koenigs et al,2008off | 0.015 | 0.032 | 25 | 0.037 | 0.014 | 25 | 4.9% | -0.88 [-1.46, -0.29] |
| Nejati (d) et al,2021 | 3.42 | 0.92 | 22 | 4.05 | 1.29 | 22 | 4.9% | -0.55 [-1.16, 0.05] |
| Nejati et al,2021 | 2.69 | 2.69 | 16 | 3.95 | 2.91 | 16 | 4.6% | -0.44 [-1.14, 0.26] |
| Weigand et al,2021off | 47.35 | 14.91 | 23 | 50.25 | 11.94 | 23 | 5.0% | -0.21 [-0.79, 0.37] |
| Tremoliere et al,2018 | 94.12 | 16.35 | 34 | 97.06 | 11.94 | 34 | 5.2% | -0.20 [-0.68, 0.27] |
| Gomez et al,2011off | 5.35 | 0.46 | 16 | 5.45 | 0.52 | 16 | 4.6% | -0.20 [-0.89, 0.50] |
| Ayache et al, 2016off | 88.4 | 15 | 16 | 90.2 | 14.6 | 16 | 4.6% | -0.12 [-0.81, 0.58] |
| Balzarotti&Colombo,2016 | 0.24 | 0.4116 | 14 | 0.25 | 0.3742 | 14 | 4.5% | -0.02 [-0.77, 0.72] |
| Nord et al,2017 | 92.54 | 5.011 | 31 | 91.5 | 5.62 | 31 | 5.2% | 0.19 [-0.31, 0.69] |
| Marie Anne et al,2016off | 88.25 | 6.69 | 35 | 86.67 | 7.03 | 35 | 5.2% | 0.23 [-0.24, 0.70] |
| Maeoka et al,2012off | 5.18 | 0.29 | 15 | 5.12 | 0.16 | 15 | 4.6% | 0.25 [-0.47, 0.97] |
| Qiao et al,2020 | 7.36 | 5.05 | 14 | 6.18 | 3.54 | 15 | 4.5% | 0.26 [-0.47, 1.00] |
| Brennan et al,2016 | 63.77 | 10.4648 | 20 | 60.35 | 8.5865 | 20 | 4.8% | 0.35 [-0.27, 0.98] |
| Boggio et al,2006 off | 10.67 | 7.05 | 26 | 8.71 | 2.98 | 26 | 5.0% | 0.36 [-0.19, 0.90] |
| Schroeder et al,2015 | 0.96 | 0.0938 | 22 | 0.91 | 0.0938 | 22 | 4.9% | 0.52 [-0.08, 1.13] |
| Nejati(c) et al,2021 | 26.1 | 2.38 | 16 | 24.3 | 2.97 | 16 | 4.6% | 0.65 [-0.06, 1.37] |
| Gilam et al, 2018off | 61 | 36.87 | 25 | 29.67 | 40.42 | 25 | 5.0% | 0.80 [0.22, 1.37] |
| Herrmann et al, 2016off | 0.12 | 0.56 | 14 | -0.49 | 0.32 | 12 | 4.2% | 1.27 [0.41, 2.13] |
| Donaldson et al, 2019off | 0.647 | 0.331 | 17 | -0.618 | 0.397 | 18 | 3.6% | 3.37 [2.30, 4.44] |
| **Total (95% CI)** | | | **470** | | | **471** | **100.0%** | **0.11 [-0.22, 0.43]** |

Heterogeneity: Tau² = 0.46; Chi² = 115.17, df = 20 (P < 0.00001); I² = 83%
Test for overall effect: Z = 0.65 (P = 0.52)

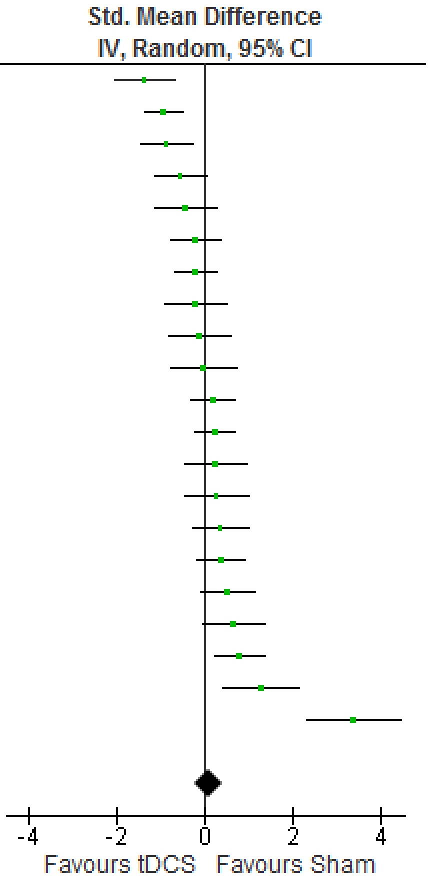


**Fig. S2** The meta-analysis of the anodal tDCS effect on emotion accuracy.

| | tDCS | | | Sham | | | | Std. Mean Difference |
|---|---|---|---|---|---|---|---|---|
| Study or Subgroup | Mean | SD | Total | Mean | SD | Total | Weight | IV, Random, 95% CI |
| Donaldson et al, 2019 ca | -238.79 | 114.339 | 18 | -57.727 | 72.658 | 18 | 19.4% | -1.85 [-2.64, -1.05] |
| JohnsonandDurrant, 2018of | 385.27 | 7.59 | 18 | 408.14 | 17.63 | 18 | 19.5% | -1.65 [-2.42, -0.88] |
| Sagliano et al,2017off | 320.42 | 25.75 | 20 | 321.72 | 29.29 | 20 | 20.4% | -0.05 [-0.67, 0.57] |
| Wolkenstein etal,2014off | 1.265 | 0.514 | 28 | 1.202 | 0.277 | 28 | 20.8% | 0.15 [-0.37, 0.68] |
| JohnsonandDurrant,2018off | 426.38 | 17.9 | 18 | 408.14 | 17.63 | 18 | 19.9% | 1.00 [0.31, 1.70] |
| **Total (95% CI)** | | | **102** | | | **102** | **100.0%** | **-0.46 [-1.44, 0.53]** |

Heterogeneity: Tau² = 1.13; Chi² = 43.28, df = 4 (P < 0.00001); I² = 91%
Test for overall effect: Z = 0.91 (P = 0.36)

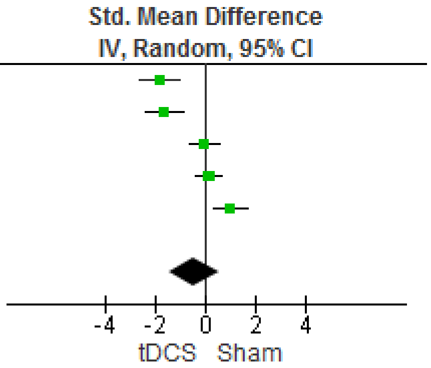


**Fig. S3** The meta-analysis of the cathodal tDCS effect on emotion RT.

| | tDCS | | | Sham | | | | Std. Mean Difference |
|---|---|---|---|---|---|---|---|---|
| Study or Subgroup | Mean | SD | Total | Mean | SD | Total | Weight | IV, Random, 95% CI |
| Koenigs et al,2008off | 0.021 | 0.045 | 25 | 0.037 | 0.014 | 25 | 17.5% | -0.47 [-1.04, 0.09] |
| Gomez et al,2011off | 5.12 | 0.15 | 16 | 5.12 | 0.4 | 16 | 16.5% | 0.00 [-0.69, 0.69] |
| Donaldson et al, 2019 ca | -0.5 | 0.336 | 18 | -0.618 | 0.397 | 18 | 16.8% | 0.31 [-0.34, 0.97] |
| Wolkenstein etal,2014off | 0.948 | 0.084 | 28 | 0.912 | 0.098 | 28 | 17.7% | 0.39 [-0.14, 0.92] |
| JohnsonandDurrant, 2018of | 0.02 | 0.03 | 18 | -0.05 | 0.05 | 18 | 15.9% | 1.66 [0.89, 2.43] |
| JohnsonandDurrant,2018off | 0.04 | 0.04 | 18 | -0.05 | 0.05 | 18 | 15.6% | 1.94 [1.14, 2.75] |
| **Total (95% CI)** | | | **123** | | | **123** | **100.0%** | **0.61 [-0.10, 1.31]** |

Heterogeneity: Tau² = 0.66; Chi² = 34.78, df = 5 (P < 0.00001); I² = 86%
Test for overall effect: Z = 1.68 (P = 0.09)

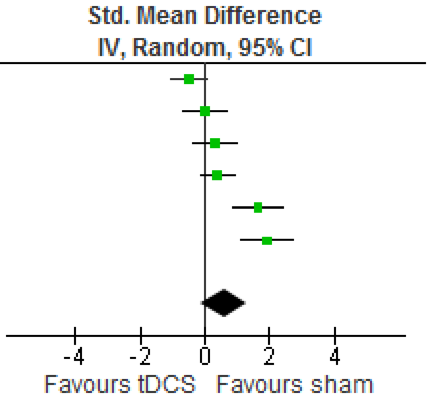


**Fig. S4** The meta-analysis of cathodal tDCS effect on emotion accuracy.

| | tDCS | | | sham | | | | Std. Mean Difference |
|---|---|---|---|---|---|---|---|---|
| Study or Subgroup | Mean | SD | Total | Mean | SD | Total | Weight | IV, Random, 95% CI |
| **3.7.1 Right frontal** | | | | | | | | |
| Jafari et al,2021 2maoff | 427.16 | 46.45 | 13 | 569.58 | 45.11 | 14 | 2.6% | -3.02 [-4.17, -1.87] |
| Jafari et al,2021off | 495.08 | 42.27 | 14 | 569.58 | 45.11 | 14 | 3.2% | -1.65 [-2.53, -0.78] |
| Colombo et al,2016 | 4.18 | 3.1307 | 10 | 6.05 | 3.1307 | 10 | 3.2% | -0.57 [-1.47, 0.33] |
| Molero-Chamizo et al,2018 | 364.94 | 175.5697 | 10 | 404.87 | 166.3358 | 10 | 3.2% | -0.22 [-1.10, 0.66] |
| Tremoliere et al,2018 | 13,000 | 6,392 | 34 | 14,693 | 9,599 | 34 | 4.2% | -0.21 [-0.68, 0.27] |
| Ayache et al, 2016off | 742.3 | 99.7 | 16 | 766.1 | 130.3 | 16 | 3.7% | -0.20 [-0.89, 0.49] |
| Sarkar et al,2014 | 1,022.22 | 296.3 | 25 | 1,070.37 | 277.8 | 25 | 4.0% | -0.17 [-0.72, 0.39] |
| Clarke et al,2020 off | 972.21 | 50.33 | 79 | 979 | 50.13 | 79 | 4.6% | -0.13 [-0.45, 0.18] |
| Dambacher et al,2015 | 3.84 | 1.16 | 16 | 4 | 1.33 | 16 | 3.7% | -0.12 [-0.82, 0.57] |
| Clarke et al,2020 benign | 949.68 | 52.14 | 79 | 955.82 | 52.04 | 79 | 4.6% | -0.12 [-0.43, 0.19] |
| Conson et al,2015 | 616.69 | 150.4 | 16 | 627.82 | 151.56 | 16 | 3.7% | -0.07 [-0.77, 0.62] |
| Sanchez Lopez et al,2016R | 340 | 54 | 27 | 342 | 69 | 27 | 4.1% | -0.03 [-0.57, 0.50] |
| Sanchezlopez etal,2018of | 390 | 94 | 27 | 391 | 79 | 27 | 4.1% | -0.01 [-0.54, 0.52] |
| Nord et al,2017 | 479.59 | 56.0674 | 31 | 477.19 | 53.3949 | 31 | 4.2% | 0.04 [-0.45, 0.54] |
| Boggio et al,2006 off | 209.15 | 77.62 | 26 | 205.45 | 61.62 | 26 | 4.1% | 0.05 [-0.49, 0.60] |
| Sagliano et al,2017off | 319.13 | 27.45 | 20 | 317.12 | 28.97 | 20 | 3.9% | 0.07 [-0.55, 0.69] |
| Marie Anne et al,2016off | 784.79 | 225.1 | 35 | 768.59 | 222.39 | 35 | 4.2% | 0.07 [-0.40, 0.54] |
| Kuehne et al,2019off | 723.05 | 72.29 | 18 | 700.9 | 58.84 | 18 | 3.8% | 0.33 [-0.33, 0.99] |
| Brennan et al,2016 | 63.8 | 10.24 | 20 | 60.35 | 8.7 | 20 | 3.9% | 0.36 [-0.27, 0.98] |
| Sanchez et al,2016off | 379.16 | 71.72 | 30 | 348.92 | 63.26 | 30 | 4.1% | 0.44 [-0.07, 0.95] |
| Okano et al,2013 | 751.4 | 71.5 | 10 | 723.7 | 45 | 10 | 3.2% | 0.44 [-0.45, 1.33] |
| Faehling and Plewnig,2016 | 1,170.09 | 45.57 | 21 | 1,110.98 | 34.62 | 22 | 3.7% | 1.44 [0.76, 2.12] |
| Fan et al,2020 | 2,374.777 | 437.793 | 18 | 1,448.777 | 124.477 | 18 | 3.1% | 2.81 [1.86, 3.76] |
| **Subtotal (95% CI)** | | | **595** | | | **597** | **87.1%** | **0.01 [-0.25, 0.28]** |
| Heterogeneity: Tau² = 0.30; Chi² = 101.90, df = 22 (P < 0.00001); I² = 78% | | | | | | | | |
| Test for overall effect: Z = 0.09 (P = 0.93) | | | | | | | | |
| **3.7.3 CentralTemporal** | | | | | | | | |
| Donaldson et al, 2019off | -618.944 | 236.218 | 17 | -57.727 | 72.658 | 18 | 2.9% | -3.18 [-4.21, -2.15] |
| Molero-Chamizo et al,2018 | 364.94 | 175.5697 | 10 | 404.87 | 166.3358 | 10 | 3.2% | -0.22 [-1.10, 0.66] |
| Qiao et al,2020 | 1,269.45 | 942.31 | 14 | 1,073.51 | 657.13 | 15 | 3.6% | 0.24 [-0.50, 0.97] |
| Okano et al,2013 | 751.4 | 71.5 | 10 | 723.7 | 45 | 10 | 3.2% | 0.44 [-0.45, 1.33] |
| **Subtotal (95% CI)** | | | **51** | | | **53** | **12.9%** | **-0.65 [-2.13, 0.82]** |
| Heterogeneity: Tau² = 2.07; Chi² = 34.37, df = 3 (P < 0.00001); I² = 91% | | | | | | | | |
| Test for overall effect: Z = 0.87 (P = 0.39) | | | | | | | | |
| **Total (95% CI)** | | | **646** | | | **650** | **100.0%** | **-0.07 [-0.34, 0.20]** |
| Heterogeneity: Tau² = 0.40; Chi² = 139.85, df = 26 (P < 0.00001); I² = 81% | | | | | | | | |
| Test for overall effect: Z = 0.49 (P = 0.62) | | | | | | | | |
| Test for subgroup differences: Chi² = 0.76, df = 1 (P = 0.38), I² = 0% | | | | | | | | |

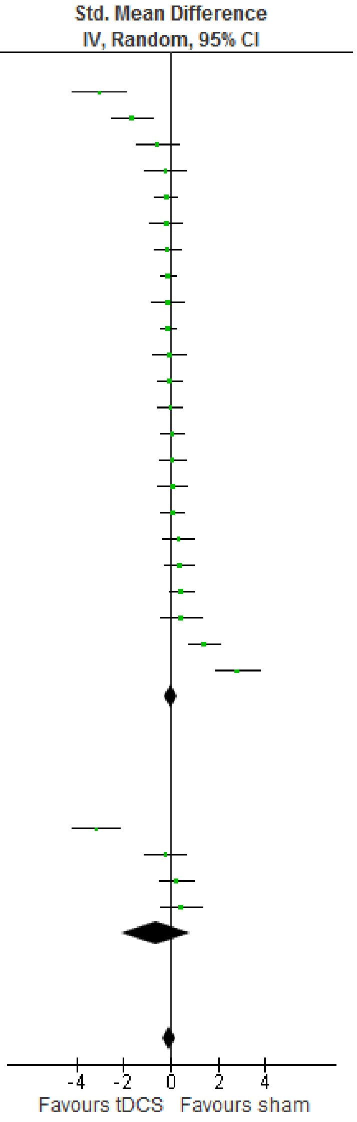


**Fig. S5** The subgroup meta-analysis based on the influence of stimulation site of tDCS on emotion RT.

| Study or Subgroup | tDCS Mean | tDCS SD | tDCS Total | sham Mean | sham SD | sham Total | Weight | Std. Mean Difference IV, Random, 95% CI |
|---|---|---|---|---|---|---|---|---|
| **3.8.1 Frontal** | | | | | | | | |
| Faehling and Plewnig,2016 | 79.29 | 1.72 | 21 | 81.7 | 1.74 | 22 | 5.2% | -1.37 [-2.04, -0.70] |
| Feeser et al, 2013off | 0.26 | 0.27 | 48 | 0.56 | 0.36 | 48 | 5.9% | -0.94 [-1.36, -0.51] |
| Nejati (d) et al,2021 | 3.42 | 0.92 | 22 | 4.05 | 1.29 | 22 | 5.4% | -0.55 [-1.16, 0.05] |
| Tremoliere et al,2018 | 94.12 | 16.35 | 34 | 97.06 | 11.94 | 34 | 5.8% | -0.20 [-0.68, 0.27] |
| Gomez et al,2011off | 5.35 | 0.46 | 16 | 5.45 | 0.52 | 16 | 5.1% | -0.20 [-0.89, 0.50] |
| Balzarotti&Colombo,2016 | 0.24 | 0.4116 | 14 | 0.25 | 0.3742 | 14 | 5.0% | -0.02 [-0.77, 0.72] |
| Nord et al,2017 | 92.54 | 5.011 | 31 | 91.5 | 5.6234 | 31 | 5.7% | 0.19 [-0.31, 0.69] |
| Marie Anne et al,2016off | 88.25 | 6.69 | 35 | 86.67 | 7.03 | 35 | 5.8% | 0.23 [-0.24, 0.70] |
| Maeoka et al,2012off | 5.18 | 0.29 | 15 | 5.12 | 0.16 | 15 | 5.1% | 0.25 [-0.47, 0.97] |
| Brennan et al,2016 | 63.77 | 10.4648 | 20 | 60.35 | 8.5865 | 20 | 5.3% | 0.35 [-0.27, 0.98] |
| Boggio et al,2006 off | 10.67 | 7.05 | 26 | 8.71 | 2.98 | 26 | 5.6% | 0.36 [-0.19, 0.90] |
| Schroeder et al,2015 | 0.96 | 0.0938 | 22 | 0.91 | 0.0938 | 22 | 5.4% | 0.52 [-0.08, 1.13] |
| Nejati(c) et al,2021 | 26.1 | 2.38 | 16 | 24.3 | 2.97 | 16 | 5.1% | 0.65 [-0.06, 1.37] |
| Herrmann et al, 2016off | 0.12 | 0.56 | 14 | -0.49 | 0.32 | 12 | 4.6% | 1.27 [0.41, 2.13] |
| **Subtotal (95% CI)** | | | **334** | | | **333** | **75.0%** | **0.01 [-0.32, 0.35]** |
| Heterogeneity: Tau² = 0.31; Chi² = 57.85, df = 13 (P < 0.00001); I² = 78% | | | | | | | | |
| Test for overall effect: Z = 0.09 (P = 0.93) | | | | | | | | |
| **3.8.3 CentralTemporal** | | | | | | | | |
| Nejati et al,2021 | 2.69 | 2.69 | 16 | 3.95 | 2.91 | 16 | 5.1% | -0.44 [-1.14, 0.26] |
| Weigand et al,2021off | 47.35 | 14.91 | 23 | 50.25 | 11.94 | 23 | 5.5% | -0.21 [-0.79, 0.37] |
| Qiao et al,2020 | 7.36 | 5.05 | 14 | 6.18 | 3.54 | 15 | 5.0% | 0.26 [-0.47, 1.00] |
| Gilam et al, 2018off | 61 | 36.87 | 25 | 29.67 | 40.42 | 25 | 5.5% | 0.80 [0.22, 1.37] |
| Donaldson et al, 2019off | 0.647 | 0.331 | 17 | -0.618 | 0.397 | 18 | 4.0% | 3.37 [2.30, 4.44] |
| **Subtotal (95% CI)** | | | **95** | | | **97** | **25.0%** | **0.69 [-0.31, 1.70]** |
| Heterogeneity: Tau² = 1.17; Chi² = 41.39, df = 4 (P < 0.00001); I² = 90% | | | | | | | | |
| Test for overall effect: Z = 1.35 (P = 0.18) | | | | | | | | |
| **Total (95% CI)** | | | **429** | | | **430** | **100.0%** | **0.17 [-0.17, 0.52]** |
| Heterogeneity: Tau² = 0.47; Chi² = 106.12, df = 18 (P < 0.00001); I² = 83% | | | | | | | | |
| Test for overall effect: Z = 0.99 (P = 0.32) | | | | | | | | |
| Test for subgroup differences: Chi² = 1.56, df = 1 (P = 0.21), I² = 36.0% | | | | | | | | |

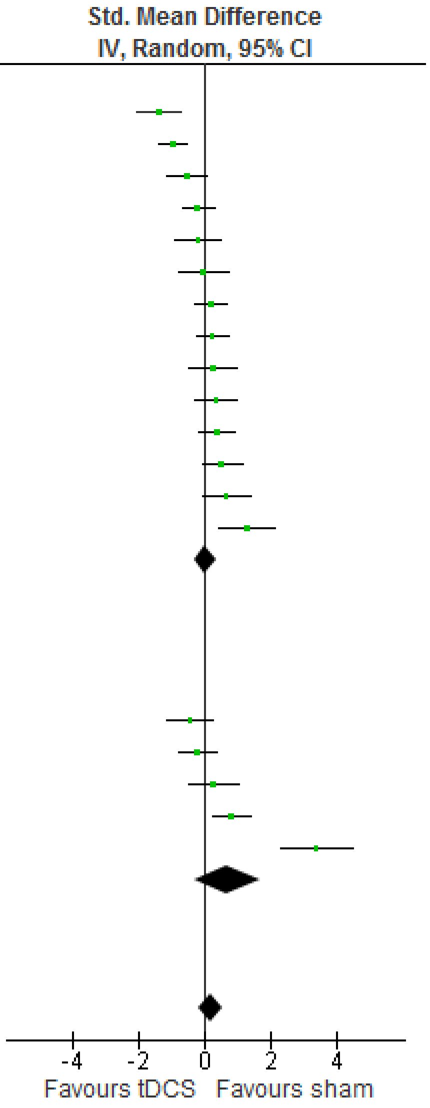


**Fig. S6** The subgroup meta-analysis based on the influence of stimulation site of tDCS on emotion accuracy.

| Study or Subgroup | tDCS Mean | tDCS SD | tDCS Total | sham Mean | sham SD | sham Total | Weight | Std. Mean Difference IV, Random, 95% CI |
|---|---|---|---|---|---|---|---|---|
| **3.13.1 >20 min** | | | | | | | | |
| Sarkar et al,2014 | 1,022.22 | 296.3 | 25 | 1,070.37 | 277.8 | 25 | 4.8% | -0.17 [-0.72, 0.39] |
| Brennan et al,2016 | 63.8 | 10.24 | 20 | 60.35 | 8.7 | 20 | 4.6% | 0.36 [-0.27, 0.98] |
| Faehling and Plewnig,2016 | 1,170.09 | 45.57 | 21 | 1,110.98 | 34.62 | 22 | 4.5% | 1.44 [0.76, 2.12] |
| **Subtotal (95% CI)** | | | **66** | | | **67** | **14.0%** | **0.53 [-0.38, 1.44]** |
| Heterogeneity: Tau² = 0.54; Chi² = 12.97, df = 2 (P = 0.002); I² = 85% | | | | | | | | |
| Test for overall effect: Z = 1.14 (P = 0.26) | | | | | | | | |
| **3.13.2 15 min** | | | | | | | | |
| Conson et al,2015 | 616.69 | 150.4 | 16 | 627.82 | 151.56 | 16 | 4.5% | -0.07 [-0.77, 0.62] |
| Sagliano et al,2017off | 319.13 | 27.45 | 20 | 317.12 | 28.97 | 20 | 4.7% | 0.07 [-0.55, 0.69] |
| **Subtotal (95% CI)** | | | **36** | | | **36** | **9.1%** | **0.01 [-0.46, 0.47]** |
| Heterogeneity: Tau² = 0.00; Chi² = 0.09, df = 1 (P = 0.77); I² = 0% | | | | | | | | |
| Test for overall effect: Z = 0.03 (P = 0.98) | | | | | | | | |
| **3.13.3 20 min** | | | | | | | | |
| Donaldson et al, 2019off | -618.944 | 236.218 | 17 | -57.727 | 72.658 | 18 | 3.5% | -3.18 [-4.21, -2.15] |
| Jafari et al,2021 2maoff | 427.16 | 46.45 | 13 | 569.58 | 45.11 | 14 | 3.2% | -3.02 [-4.17, -1.87] |
| Jafari et al,2021off | 495.08 | 42.27 | 14 | 569.58 | 45.11 | 14 | 3.9% | -1.65 [-2.53, -0.78] |
| Colombo et al,2016 | 4.18 | 3.1307 | 10 | 6.05 | 3.1307 | 10 | 3.9% | -0.57 [-1.47, 0.33] |
| Tremoliere et al,2018 | 13,000 | 6,392 | 34 | 14,693 | 9,599 | 34 | 5.0% | -0.21 [-0.68, 0.27] |
| Ayache et al, 2016off | 742.3 | 99.7 | 16 | 766.1 | 130.3 | 16 | 4.4% | -0.20 [-0.89, 0.49] |
| Clarke et al,2020 off | 972.21 | 50.33 | 79 | 979 | 50.13 | 79 | 5.4% | -0.13 [-0.45, 0.18] |
| Dambacher et al,2015 | 3.84 | 1.16 | 16 | 4 | 1.33 | 16 | 4.5% | -0.12 [-0.82, 0.57] |
| Clarke et al,2020 benign | 949.68 | 52.14 | 79 | 955.82 | 52.04 | 79 | 5.4% | -0.12 [-0.43, 0.19] |
| Sanchez Lopez et al,2016R | 340 | 54 | 27 | 342 | 69 | 27 | 4.9% | -0.03 [-0.57, 0.50] |
| Sanchezlopez etal,2018of | 390 | 94 | 27 | 391 | 79 | 27 | 4.9% | -0.01 [-0.54, 0.52] |
| Nord et al,2017 | 479.59 | 56.0674 | 31 | 477.19 | 53.3949 | 31 | 5.0% | 0.04 [-0.45, 0.54] |
| Boggio et al,2006 off | 209.15 | 77.62 | 26 | 205.45 | 61.62 | 26 | 4.9% | 0.05 [-0.49, 0.60] |
| Marie Anne et al,2016off | 784.79 | 225.1 | 35 | 768.59 | 222.39 | 35 | 5.0% | 0.07 [-0.40, 0.54] |
| Qiao et al,2020 | 1,269.45 | 942.31 | 14 | 1,073.51 | 657.13 | 15 | 4.3% | 0.24 [-0.50, 0.97] |
| Sanchez et al,2016off | 379.16 | 71.72 | 30 | 348.92 | 63.26 | 30 | 4.9% | 0.44 [-0.07, 0.95] |
| Fan et al,2020 | 2,374.777 | 437.793 | 18 | 1,448.777 | 124.477 | 18 | 3.7% | 2.81 [1.86, 3.76] |
| **Subtotal (95% CI)** | | | **486** | | | **489** | **76.9%** | **-0.25 [-0.62, 0.12]** |
| Heterogeneity: Tau² = 0.48; Chi² = 114.88, df = 16 (P < 0.00001); I² = 86% | | | | | | | | |
| Test for overall effect: Z = 1.34 (P = 0.18) | | | | | | | | |
| **Total (95% CI)** | | | **588** | | | **592** | **100.0%** | **-0.12 [-0.43, 0.20]** |
| Heterogeneity: Tau² = 0.45; Chi² = 136.15, df = 21 (P < 0.00001); I² = 85% | | | | | | | | |
| Test for overall effect: Z = 0.74 (P = 0.46) | | | | | | | | |
| Test for subgroup differences: Chi² = 2.69, df = 2 (P = 0.26), I² = 25.6% | | | | | | | | |

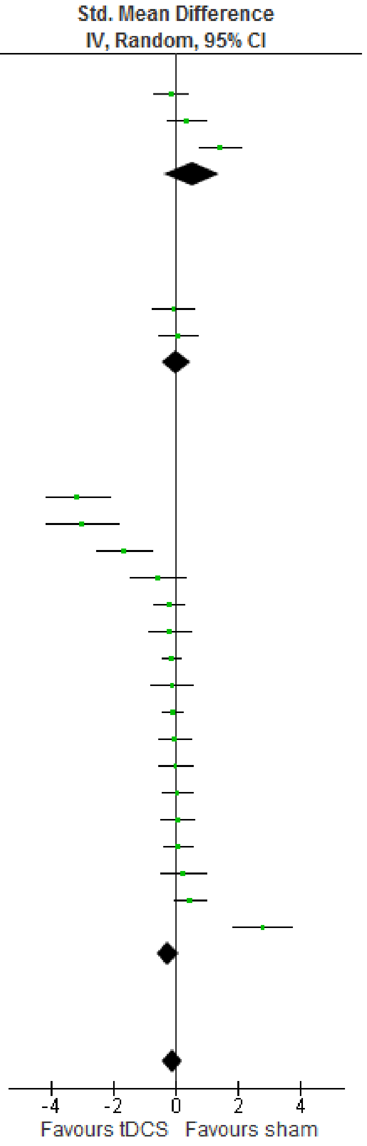


**Fig. S7** The subgroup meta-analysis based on the influence of duration stimulation of tDCS on emotion RT.

| Study or Subgroup | tDCS Mean | tDCS SD | tDCS Total | sham Mean | sham SD | sham Total | Weight | Std. Mean Difference IV, Random, 95% CI |
|---|---|---|---|---|---|---|---|---|
| **3.14.1 >20 min** | | | | | | | | |
| Faehling and Plewnig,2016 | 79.29 | 1.72 | 21 | 81.7 | 1.74 | 22 | 4.7% | -1.37 [-2.04, -0.70] |
| Koenigs et al,2008off | 0.015 | 0.032 | 25 | 0.037 | 0.014 | 25 | 4.9% | -0.88 [-1.46, -0.29] |
| Brennan et al,2016 | 63.77 | 10.4648 | 20 | 60.35 | 8.5865 | 20 | 4.8% | 0.35 [-0.27, 0.98] |
| Gilam et al, 2018off | 61 | 36.87 | 25 | 29.67 | 40.42 | 25 | 5.0% | 0.80 [0.22, 1.37] |
| **Subtotal (95% CI)** | | | **91** | | | **92** | **19.4%** | **-0.27 [-1.26, 0.72]** |
| Heterogeneity: Tau² = 0.92; Chi² = 31.23, df = 3 (P < 0.00001); I² = 90% | | | | | | | | |
| Test for overall effect: Z = 0.53 (P = 0.59) | | | | | | | | |
| **3.14.2 15 min** | | | | | | | | |
| Nejati (d) et al,2021 | 3.42 | 0.92 | 22 | 4.05 | 1.29 | 22 | 4.9% | -0.55 [-1.16, 0.05] |
| Nejati et al,2021 | 2.69 | 2.69 | 16 | 3.95 | 2.91 | 16 | 4.6% | -0.44 [-1.14, 0.26] |
| Balzarotti&Colombo,2016 | 0.24 | 0.4116 | 14 | 0.25 | 0.3742 | 14 | 4.5% | -0.02 [-0.77, 0.72] |
| Nejati(c) et al,2021 | 26.1 | 2.38 | 16 | 24.3 | 2.97 | 16 | 4.6% | 0.65 [-0.06, 1.37] |
| **Subtotal (95% CI)** | | | **68** | | | **68** | **18.6%** | **-0.11 [-0.64, 0.43]** |
| Heterogeneity: Tau² = 0.18; Chi² = 7.31, df = 3 (P = 0.06); I² = 59% | | | | | | | | |
| Test for overall effect: Z = 0.39 (P = 0.69) | | | | | | | | |
| **3.14.3 20min** | | | | | | | | |
| Feeser et al, 2013off | 0.26 | 0.27 | 48 | 0.56 | 0.36 | 48 | 5.4% | -0.94 [-1.36, -0.51] |
| Weigand et al,2021off | 47.35 | 14.91 | 23 | 50.25 | 11.94 | 23 | 5.0% | -0.21 [-0.79, 0.37] |
| Tremoliere et al,2018 | 94.12 | 16.35 | 34 | 97.06 | 11.94 | 34 | 5.2% | -0.20 [-0.68, 0.27] |
| Gomez et al,2011off | 5.35 | 0.46 | 16 | 5.45 | 0.52 | 16 | 4.6% | -0.20 [-0.89, 0.50] |
| Ayache et al, 2016off | 88.4 | 15 | 16 | 90.2 | 14.6 | 16 | 4.6% | -0.12 [-0.81, 0.58] |
| Nord et al,2017 | 92.54 | 5.011 | 31 | 91.5 | 5.62 | 31 | 5.2% | 0.19 [-0.31, 0.69] |
| Marie Anne et al,2016off | 88.25 | 6.69 | 35 | 86.67 | 7.03 | 35 | 5.2% | 0.23 [-0.24, 0.70] |
| Maeoka et al,2012off | 5.18 | 0.29 | 15 | 5.12 | 0.16 | 15 | 4.6% | 0.25 [-0.47, 0.97] |
| Qiao et al,2020 | 7.36 | 5.05 | 14 | 6.18 | 3.54 | 15 | 4.5% | 0.26 [-0.47, 1.00] |
| Boggio et al,2006 off | 10.67 | 7.05 | 26 | 8.71 | 2.98 | 26 | 5.0% | 0.36 [-0.19, 0.90] |
| Schroeder et al,2015 | 0.96 | 0.0938 | 22 | 0.91 | 0.0938 | 22 | 4.9% | 0.52 [-0.08, 1.13] |
| Herrmann et al, 2016off | 0.12 | 0.56 | 14 | -0.49 | 0.32 | 12 | 4.2% | 1.27 [0.41, 2.13] |
| Donaldson et al, 2019off | 0.647 | 0.331 | 17 | -0.618 | 0.397 | 18 | 3.6% | 3.37 [2.30, 4.44] |
| **Subtotal (95% CI)** | | | **311** | | | **311** | **62.0%** | **0.29 [-0.13, 0.70]** |
| Heterogeneity: Tau² = 0.47; Chi² = 73.38, df = 12 (P < 0.00001); I² = 84% | | | | | | | | |
| Test for overall effect: Z = 1.36 (P = 0.17) | | | | | | | | |
| **Total (95% CI)** | | | **470** | | | **471** | **100.0%** | **0.11 [-0.22, 0.43]** |
| Heterogeneity: Tau² = 0.46; Chi² = 115.17, df = 20 (P < 0.00001); I² = 83% | | | | | | | | |
| Test for overall effect: Z = 0.65 (P = 0.52) | | | | | | | | |
| Test for subgroup differences: Chi² = 1.89, df = 2 (P = 0.39), I² = 0% | | | | | | | | |

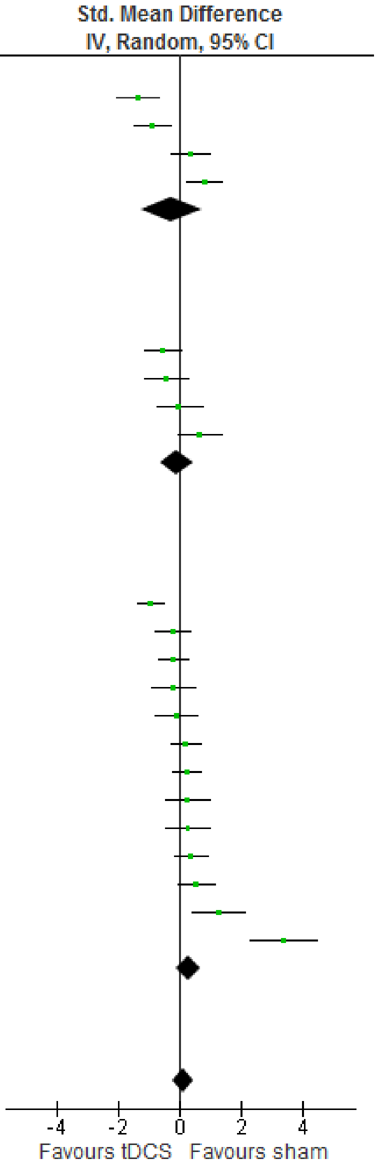


**Fig. S8** The subgroup meta-analysis based on the influence of duration stimulation of tDCS on emotion accuracy.

| Study or Subgroup | tDCS Mean | tDCS SD | tDCS Total | sham Mean | sham SD | sham Total | Weight | Std. Mean Difference IV, Random, 95% CI |
|---|---|---|---|---|---|---|---|---|
| **3.15.1 1mA** | | | | | | | | |
| Jafari et al,2021off | 495.08 | 42.27 | 14 | 569.58 | 45.11 | 14 | 3.8% | -1.65 [-2.53, -0.78] |
| Sarkar et al,2014 | 1,022.22 | 296.3 | 25 | 1,070.37 | 277.8 | 25 | 4.6% | -0.17 [-0.72, 0.39] |
| Conson et al,2015 | 616.69 | 150.4 | 16 | 627.82 | 151.56 | 16 | 4.3% | -0.07 [-0.77, 0.62] |
| Nord et al,2017 | 479.59 | 56.0674 | 31 | 477.19 | 53.3949 | 31 | 4.8% | 0.04 [-0.45, 0.54] |
| Sagliano et al,2017off | 319.13 | 27.45 | 20 | 317.12 | 28.97 | 20 | 4.5% | 0.07 [-0.55, 0.69] |
| Kuehne et al,2019off | 723.05 | 72.29 | 18 | 700.9 | 58.84 | 18 | 4.3% | 0.33 [-0.33, 0.99] |
| **Subtotal (95% CI)** | | | **124** | | | **124** | **26.2%** | **-0.18 [-0.62, 0.26]** |
| Heterogeneity: Tau² = 0.19; Chi² = 14.39, df = 5 (P = 0.01); I² = 65% | | | | | | | | |
| Test for overall effect: Z = 0.80 (P = 0.42) | | | | | | | | |
| **3.15.2 1.5 mA** | | | | | | | | |
| Colombo et al,2016 | 4.18 | 3.1307 | 10 | 6.05 | 3.1307 | 10 | 3.7% | -0.57 [-1.47, 0.33] |
| Brennan et al,2016 | 63.8 | 10.24 | 20 | 60.35 | 8.7 | 20 | 4.4% | 0.36 [-0.27, 0.98] |
| Faehling and Plewnig,2016 | 1,170.09 | 45.57 | 21 | 1,110.98 | 34.62 | 22 | 4.3% | 1.44 [0.76, 2.12] |
| **Subtotal (95% CI)** | | | **51** | | | **52** | **12.4%** | **0.44 [-0.63, 1.50]** |
| Heterogeneity: Tau² = 0.75; Chi² = 12.96, df = 2 (P = 0.002); I² = 85% | | | | | | | | |
| Test for overall effect: Z = 0.81 (P = 0.42) | | | | | | | | |
| **3.15.5 2 mA** | | | | | | | | |
| Donaldson et al, 2019off | -618.944 | 236.218 | 17 | -57.727 | 72.658 | 18 | 3.3% | -3.18 [-4.21, -2.15] |
| Jafari et al,2021 2maoff | 427.16 | 46.45 | 13 | 569.58 | 45.11 | 14 | 3.1% | -3.02 [-4.17, -1.87] |
| Tremoliere et al,2018 | 13,000 | 6,392 | 34 | 14,693 | 9,599 | 34 | 4.8% | -0.21 [-0.68, 0.27] |
| Ayache et al, 2016off | 742.3 | 99.7 | 16 | 766.1 | 130.3 | 16 | 4.2% | -0.20 [-0.89, 0.49] |
| Clarke et al,2020 off | 972.21 | 50.33 | 79 | 979 | 50.13 | 79 | 5.2% | -0.13 [-0.45, 0.18] |
| Dambacher et al,2015 | 3.84 | 1.16 | 16 | 4 | 1.33 | 16 | 4.3% | -0.12 [-0.82, 0.57] |
| Clarke et al,2020 benign | 949.68 | 52.14 | 79 | 955.82 | 52.04 | 79 | 5.2% | -0.12 [-0.43, 0.19] |
| Sanchez Lopez et al,2016R | 340 | 54 | 27 | 342 | 69 | 27 | 4.7% | -0.03 [-0.57, 0.50] |
| Sanchezlopez etal,2018of | 390 | 94 | 27 | 391 | 79 | 27 | 4.7% | -0.01 [-0.54, 0.52] |
| Boggio et al,2006 off | 209.15 | 77.62 | 26 | 205.45 | 61.62 | 26 | 4.7% | 0.05 [-0.49, 0.60] |
| Marie Anne et al,2016off | 784.79 | 225.1 | 35 | 768.59 | 222.39 | 35 | 4.8% | 0.07 [-0.40, 0.54] |
| Qiao et al,2020 | 1,269.45 | 942.31 | 14 | 1,073.51 | 657.13 | 15 | 4.1% | 0.24 [-0.50, 0.97] |
| Sanchez et al,2016off | 379.16 | 71.72 | 30 | 348.92 | 63.26 | 30 | 4.7% | 0.44 [-0.07, 0.95] |
| Fan et al,2020 | 2,374.777 | 437.793 | 18 | 1,448.777 | 124.477 | 18 | 3.6% | 2.81 [1.86, 3.76] |
| **Subtotal (95% CI)** | | | **431** | | | **434** | **61.4%** | **-0.17 [-0.58, 0.24]** |
| Heterogeneity: Tau² = 0.49; Chi² = 101.28, df = 13 (P < 0.00001); I² = 87% | | | | | | | | |
| Test for overall effect: Z = 0.82 (P = 0.41) | | | | | | | | |
| **Total (95% CI)** | | | **606** | | | **610** | **100.0%** | **-0.10 [-0.40, 0.21]** |
| Heterogeneity: Tau² = 0.44; Chi² = 137.32, df = 22 (P < 0.00001); I² = 84% | | | | | | | | |
| Test for overall effect: Z = 0.63 (P = 0.53) | | | | | | | | |
| Test for subgroup differences: Chi² = 1.18, df = 2 (P = 0.55), I² = 0% | | | | | | | | |

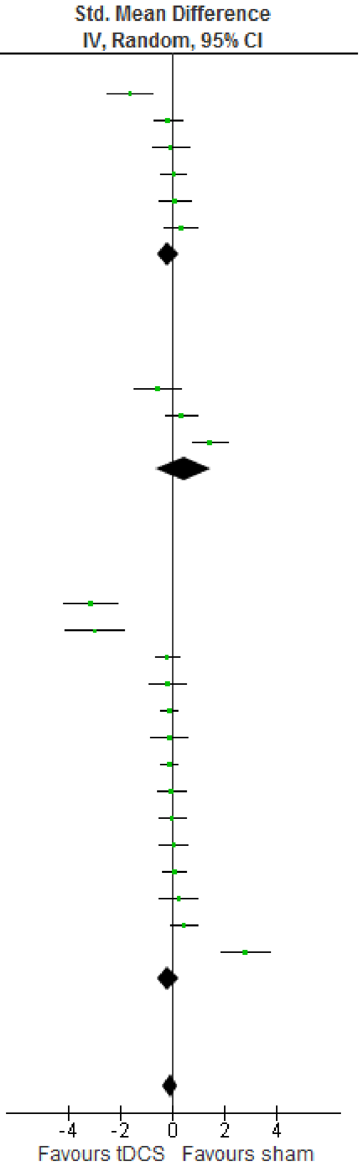


**Fig. S9** The subgroup meta-analysis based on the influence of current level of tDCS on emotion RT.

| Study or Subgroup | tDCS Mean | tDCS SD | tDCS Total | sham Mean | sham SD | sham Total | Weight | Std. Mean Difference IV, Random, 95% CI |
|---|---|---|---|---|---|---|---|---|
| **3.16.1 1mA** | | | | | | | | |
| Nejati et al,2021 | 2.69 | 2.69 | 16 | 3.95 | 2.91 | 16 | 5.1% | -0.44 [-1.14, 0.26] |
| Weigand et al,2021off | 47.35 | 14.91 | 23 | 50.25 | 11.94 | 23 | 5.5% | -0.21 [-0.79, 0.37] |
| Nord et al,2017 | 92.54 | 5.011 | 31 | 91.5 | 5.62 | 31 | 5.7% | 0.19 [-0.31, 0.69] |
| Maeoka et al,2012off | 5.18 | 0.29 | 15 | 5.12 | 0.16 | 15 | 5.0% | 0.25 [-0.47, 0.97] |
| Schroeder et al,2015 | 0.96 | 0.0938 | 22 | 0.91 | 0.0938 | 22 | 5.4% | 0.52 [-0.08, 1.13] |
| Nejati(c) et al,2021 | 26.1 | 2.38 | 16 | 24.3 | 2.97 | 16 | 5.1% | 0.65 [-0.06, 1.37] |
| **Subtotal (95% CI)** | | | **123** | | | **123** | **31.8%** | **0.16 [-0.16, 0.47]** |
| Heterogeneity: Tau² = 0.05; Chi² = 7.66, df = 5 (P = 0.18); I² = 35% | | | | | | | | |
| Test for overall effect: Z = 0.98 (P = 0.33) | | | | | | | | |
| **3.16.2 1.5mA** | | | | | | | | |
| Faehling and Plewnig,2016 | 79.29 | 1.72 | 21 | 81.7 | 1.74 | 22 | 5.2% | -1.37 [-2.04, -0.70] |
| Feeser et al, 2013off | 0.26 | 0.27 | 48 | 0.56 | 0.36 | 48 | 5.9% | -0.94 [-1.36, -0.51] |
| Nejati (d) et al,2021 | 3.42 | 0.92 | 22 | 4.05 | 1.29 | 22 | 5.4% | -0.55 [-1.16, 0.05] |
| Balzarotti&Colombo,2016 | 0.24 | 0.4116 | 14 | 0.25 | 0.3742 | 14 | 5.0% | -0.02 [-0.77, 0.72] |
| Brennan et al,2016 | 63.77 | 10.4648 | 20 | 60.35 | 8.5865 | 20 | 5.3% | 0.35 [-0.27, 0.98] |
| Gilam et al, 2018off | 61 | 36.87 | 25 | 29.67 | 40.42 | 25 | 5.5% | 0.80 [0.22, 1.37] |
| **Subtotal (95% CI)** | | | **150** | | | **151** | **32.3%** | **-0.29 [-0.95, 0.37]** |
| Heterogeneity: Tau² = 0.58; Chi² = 37.34, df = 5 (P < 0.00001); I² = 87% | | | | | | | | |
| Test for overall effect: Z = 0.87 (P = 0.38) | | | | | | | | |
| **3.16.5 2mA** | | | | | | | | |
| Tremoliere et al,2018 | 94.12 | 16.35 | 34 | 97.06 | 11.94 | 34 | 5.8% | -0.20 [-0.68, 0.27] |
| Ayache et al, 2016off | 88.4 | 15 | 16 | 90.2 | 14.6 | 16 | 5.1% | -0.12 [-0.81, 0.58] |
| Marie Anne et al,2016off | 88.25 | 6.69 | 35 | 86.67 | 7.03 | 35 | 5.8% | 0.23 [-0.24, 0.70] |
| Qiao et al,2020 | 7.36 | 5.05 | 14 | 6.18 | 3.54 | 15 | 5.0% | 0.26 [-0.47, 1.00] |
| Boggio et al,2006 off | 10.67 | 7.05 | 26 | 8.71 | 2.98 | 26 | 5.6% | 0.36 [-0.19, 0.90] |
| Herrmann et al, 2016off | 0.12 | 0.56 | 14 | -0.49 | 0.32 | 12 | 4.6% | 1.27 [0.41, 2.13] |
| Donaldson et al, 2019off | 0.647 | 0.331 | 17 | -0.618 | 0.397 | 18 | 4.0% | 3.37 [2.30, 4.44] |
| **Subtotal (95% CI)** | | | **156** | | | **156** | **35.9%** | **0.64 [-0.00, 1.28]** |
| Heterogeneity: Tau² = 0.63; Chi² = 42.35, df = 6 (P < 0.00001); I² = 86% | | | | | | | | |
| Test for overall effect: Z = 1.95 (P = 0.05) | | | | | | | | |
| **Total (95% CI)** | | | **429** | | | **430** | **100.0%** | **0.18 [-0.16, 0.52]** |
| Heterogeneity: Tau² = 0.47; Chi² = 105.87, df = 18 (P < 0.00001); I² = 83% | | | | | | | | |
| Test for overall effect: Z = 1.01 (P = 0.31) | | | | | | | | |
| Test for subgroup differences: Chi² = 3.94, df = 2 (P = 0.14), I² = 49.3% | | | | | | | | |

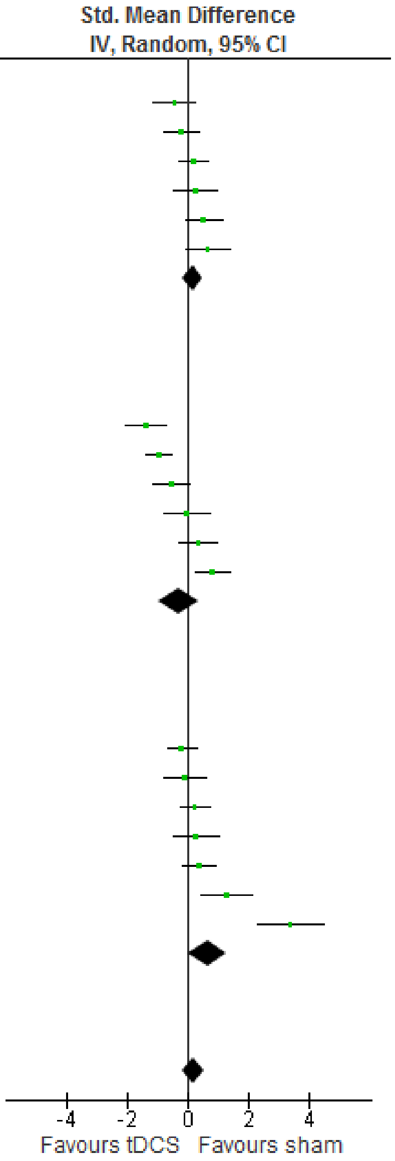


**Fig. S10** The subgroup meta-analysis based on the influence of current level of tDCS on emotion accuracy.

| Study or Subgroup | tDCS Mean | tDCS SD | tDCS Total | sham Mean | sham SD | sham Total | Weight | Std. Mean Difference IV, Random, 95% CI |
|---|---|---|---|---|---|---|---|---|
| **3.19.1 clinical** | | | | | | | | |
| Jafari et al,2021 2maoff | 427.16 | 46.45 | 13 | 569.58 | 45.11 | 14 | 3.1% | -3.02 [-4.17, -1.87] |
| Jafari et al,2021off | 495.08 | 42.27 | 14 | 569.58 | 45.11 | 14 | 3.7% | -1.65 [-2.53, -0.78] |
| Ayache et al, 2016off | 742.3 | 99.7 | 16 | 766.1 | 130.3 | 16 | 4.2% | -0.20 [-0.89, 0.49] |
| Sarkar et al,2014 | 1,022.22 | 296.3 | 25 | 1,070.37 | 277.8 | 25 | 4.5% | -0.17 [-0.72, 0.39] |
| Boggio et al,2006 off | 209.15 | 77.62 | 26 | 205.45 | 61.62 | 26 | 4.5% | 0.05 [-0.49, 0.60] |
| Qiao et al,2020 | 1,269.45 | 942.31 | 14 | 1,073.51 | 657.13 | 15 | 4.1% | 0.24 [-0.50, 0.97] |
| **Subtotal (95% CI)** | | | **108** | | | **110** | **24.1%** | **-0.69 [-1.45, 0.07]** |
| Heterogeneity: Tau² = 0.75; Chi² = 34.24, df = 5 (P < 0.00001); I² = 85% | | | | | | | | |
| Test for overall effect: Z = 1.78 (P = 0.08) | | | | | | | | |
| **3.19.2 healthy** | | | | | | | | |
| Donaldson et al, 2019off | -618.944 | 236.218 | 17 | -57.727 | 72.658 | 18 | 3.4% | -3.18 [-4.21, -2.15] |
| Colombo et al,2016 | 4.18 | 3.1307 | 10 | 6.05 | 3.1307 | 10 | 3.7% | -0.57 [-1.47, 0.33] |
| Molero-Chamizo et al,2018 | 364.94 | 175.5697 | 10 | 404.87 | 166.3358 | 10 | 3.7% | -0.22 [-1.10, 0.66] |
| Tremoliere et al,2018 | 13,000 | 6,392 | 34 | 14,693 | 9,599 | 34 | 4.7% | -0.21 [-0.68, 0.27] |
| Clarke et al,2020 off | 972.21 | 50.33 | 79 | 979 | 50.13 | 79 | 5.0% | -0.13 [-0.45, 0.18] |
| Dambacher et al,2015 | 3.84 | 1.16 | 16 | 4 | 1.33 | 16 | 4.2% | -0.12 [-0.82, 0.57] |
| Conson et al,2015 | 616.69 | 150.4 | 16 | 627.82 | 151.56 | 16 | 4.2% | -0.07 [-0.77, 0.62] |
| Sanchez Lopez et al,2016R | 340 | 54 | 27 | 342 | 69 | 27 | 4.6% | -0.03 [-0.57, 0.50] |
| Balzarotti&Colombo,2016 | 0.24 | 0.4116 | 14 | 0.25 | 0.3742 | 14 | 4.1% | -0.02 [-0.77, 0.72] |
| Sanchezlopez etal,2018of | 390 | 94 | 27 | 391 | 79 | 27 | 4.6% | -0.01 [-0.54, 0.52] |
| Nord et al,2017 | 479.59 | 56.0674 | 31 | 477.19 | 53.3949 | 31 | 4.6% | 0.04 [-0.45, 0.54] |
| Sagliano et al,2017off | 319.13 | 27.45 | 20 | 317.12 | 28.97 | 20 | 4.4% | 0.07 [-0.55, 0.69] |
| Marie Anne et al,2016off | 784.79 | 225.1 | 35 | 768.59 | 222.39 | 35 | 4.7% | 0.07 [-0.40, 0.54] |
| Kuehne et al,2019off | 723.05 | 72.29 | 18 | 700.9 | 58.84 | 18 | 4.3% | 0.33 [-0.33, 0.99] |
| Brennan et al,2016 | 63.8 | 10.24 | 20 | 60.35 | 8.7 | 20 | 4.4% | 0.36 [-0.27, 0.98] |
| Okano et al,2013 | 751.4 | 71.5 | 10 | 723.7 | 45 | 10 | 3.7% | 0.44 [-0.45, 1.33] |
| Faehling and Plewnig,2016 | 1,170.09 | 45.57 | 21 | 1,110.98 | 34.62 | 22 | 4.2% | 1.44 [0.76, 2.12] |
| Fan et al,2020 | 2,374.777 | 437.793 | 18 | 1,448.777 | 124.477 | 18 | 3.5% | 2.81 [1.86, 3.76] |
| **Subtotal (95% CI)** | | | **423** | | | **425** | **75.9%** | **0.07 [-0.27, 0.41]** |
| Heterogeneity: Tau² = 0.42; Chi² = 93.87, df = 17 (P < 0.00001); I² = 82% | | | | | | | | |
| Test for overall effect: Z = 0.40 (P = 0.69) | | | | | | | | |
| **Total (95% CI)** | | | **531** | | | **535** | **100.0%** | **-0.11 [-0.42, 0.21]** |
| Heterogeneity: Tau² = 0.48; Chi² = 135.03, df = 23 (P < 0.00001); I² = 83% | | | | | | | | |
| Test for overall effect: Z = 0.67 (P = 0.50) | | | | | | | | |
| Test for subgroup differences: Chi² = 3.19, df = 1 (P = 0.07), I² = 68.7% | | | | | | | | |

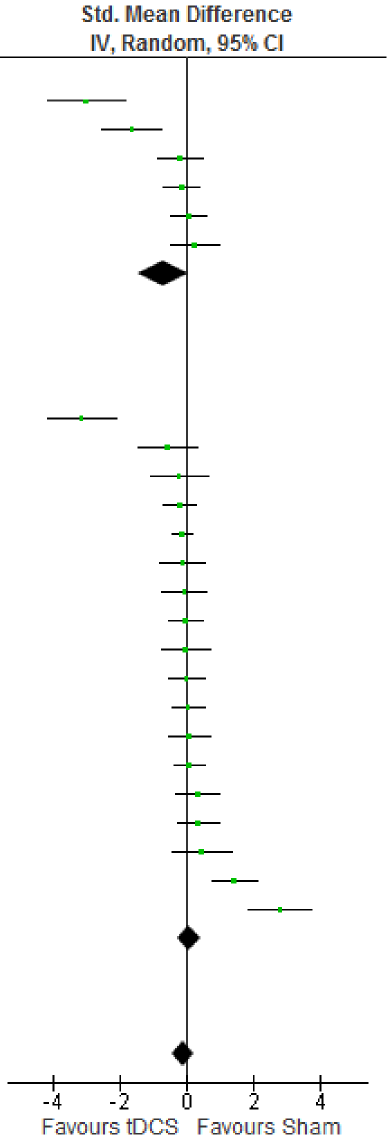


**Fig. S11** The subgroup meta-analysis based on the influence of healthy/clinical of tDCS on emotion RT

| Study or Subgroup | tDCS Mean | tDCS SD | tDCS Total | sham Mean | sham SD | sham Total | Weight | Std. Mean Difference IV, Random, 95% CI |
|---|---|---|---|---|---|---|---|---|
| **3.20.1 clinical** | | | | | | | | |
| Feeser et al, 2013off | 0.26 | 0.27 | 48 | 0.56 | 0.36 | 48 | 5.9% | -0.94 [-1.36, -0.51] |
| Nejati (d) et al,2021 | 3.42 | 0.92 | 22 | 4.05 | 1.29 | 22 | 5.4% | -0.55 [-1.16, 0.05] |
| Ayache et al, 2016off | 88.4 | 15 | 16 | 90.2 | 14.6 | 16 | 5.1% | -0.12 [-0.81, 0.58] |
| Qiao et al,2020 | 7.36 | 5.05 | 14 | 6.18 | 3.54 | 15 | 5.0% | 0.26 [-0.47, 1.00] |
| Boggio et al,2006 off | 10.67 | 7.05 | 26 | 8.71 | 2.98 | 26 | 5.6% | 0.36 [-0.19, 0.90] |
| **Subtotal (95% CI)** | | | **126** | | | **127** | **27.0%** | **-0.22 [-0.77, 0.32]** |
| Heterogeneity: Tau² = 0.29; Chi² = 17.33, df = 4 (P = 0.002); I² = 77% | | | | | | | | |
| Test for overall effect: Z = 0.81 (P = 0.42) | | | | | | | | |
| **3.20.2 healthy** | | | | | | | | |
| Faehling and Plewnig,2016 | 79.29 | 1.72 | 21 | 81.7 | 1.74 | 22 | 5.2% | -1.37 [-2.04, -0.70] |
| Nejati et al,2021 | 2.69 | 2.69 | 16 | 3.95 | 2.91 | 16 | 5.1% | -0.44 [-1.14, 0.26] |
| Weigand et al,2021off | 47.35 | 14.91 | 23 | 50.25 | 11.94 | 23 | 5.5% | -0.21 [-0.79, 0.37] |
| Tremoliere et al,2018 | 94.12 | 16.35 | 34 | 97.06 | 11.94 | 34 | 5.8% | -0.20 [-0.68, 0.27] |
| Gomez et al,2011off | 5.35 | 0.46 | 16 | 5.45 | 0.52 | 16 | 5.1% | -0.20 [-0.89, 0.50] |
| Nord et al,2017 | 92.54 | 5.011 | 31 | 91.5 | 5.6234 | 31 | 5.7% | 0.19 [-0.31, 0.69] |
| Marie Anne et al,2016off | 88.25 | 6.69 | 35 | 86.67 | 7.03 | 35 | 5.8% | 0.23 [-0.24, 0.70] |
| Maeoka et al,2012off | 5.18 | 0.29 | 15 | 5.12 | 0.16 | 15 | 5.0% | 0.25 [-0.47, 0.97] |
| Brennan et al,2016 | 63.77 | 10.4648 | 20 | 60.35 | 8.5865 | 20 | 5.3% | 0.35 [-0.27, 0.98] |
| Schroeder et al,2015 | 0.96 | 0.0938 | 22 | 0.91 | 0.0938 | 22 | 5.4% | 0.52 [-0.08, 1.13] |
| Nejati(c) et al,2021 | 26.1 | 2.38 | 16 | 24.3 | 2.97 | 16 | 5.1% | 0.65 [-0.06, 1.37] |
| Gilam et al, 2018off | 61 | 36.87 | 25 | 29.67 | 40.42 | 25 | 5.5% | 0.80 [0.22, 1.37] |
| Herrmann et al, 2016off | 0.12 | 0.56 | 14 | -0.49 | 0.32 | 12 | 4.6% | 1.27 [0.41, 2.13] |
| Donaldson et al, 2019off | 0.647 | 0.331 | 17 | -0.618 | 0.397 | 18 | 4.0% | 3.37 [2.30, 4.44] |
| **Subtotal (95% CI)** | | | **305** | | | **305** | **73.0%** | **0.31 [-0.10, 0.72]** |
| Heterogeneity: Tau² = 0.49; Chi² = 76.92, df = 13 (P < 0.00001); I² = 83% | | | | | | | | |
| Test for overall effect: Z = 1.49 (P = 0.14) | | | | | | | | |
| **Total (95% CI)** | | | **431** | | | **432** | **100.0%** | **0.17 [-0.17, 0.51]** |
| Heterogeneity: Tau² = 0.47; Chi² = 106.28, df = 18 (P < 0.00001); I² = 83% | | | | | | | | |
| Test for overall effect: Z = 0.96 (P = 0.34) | | | | | | | | |
| Test for subgroup differences: Chi² = 2.38, df = 1 (P = 0.12), I² = 58.0% | | | | | | | | |

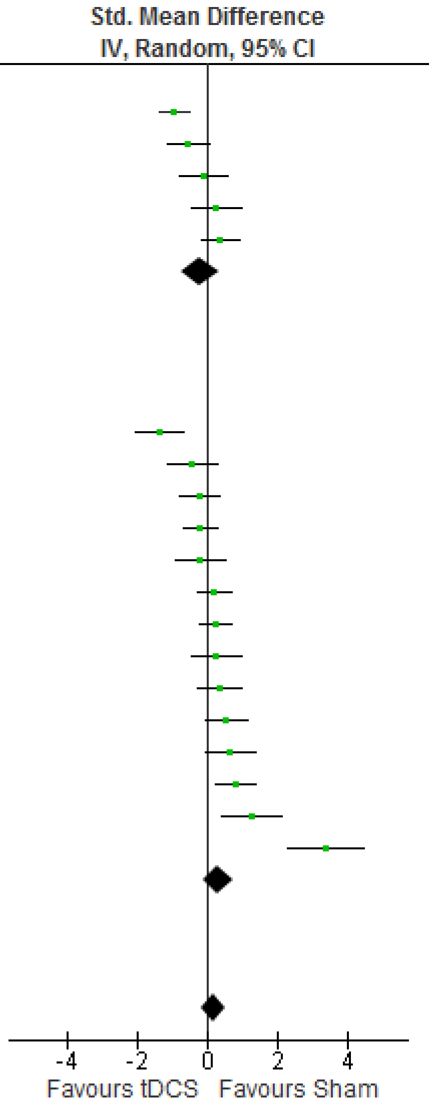


**Fig. S12** The subgroup meta-analysis based on the influence of healthy/clinical of tDCS on emotion accuracy.

| Study or Subgroup | tDCS Mean | tDCS SD | tDCS Total | sham Mean | sham SD | sham Total | Weight | Std. Mean Difference IV, Random, 95% CI |
|---|---|---|---|---|---|---|---|---|
| **3.17.1 on RT** | | | | | | | | |
| Colombo et al,2016 | 4.18 | 3.1307 | 10 | 6.05 | 3.1307 | 10 | 3.4% | -0.57 [-1.47, 0.33] |
| Molero-Chamizo et al,2018 | 364.94 | 175.5697 | 10 | 404.87 | 166.3358 | 10 | 3.5% | -0.22 [-1.10, 0.66] |
| Tremoliere et al,2018 | 13,000 | 6,392 | 34 | 14,693 | 9,599 | 34 | 4.5% | -0.21 [-0.68, 0.27] |
| Sarkar et al,2014 | 1,022.22 | 296.3 | 25 | 1,070.37 | 277.8 | 25 | 4.3% | -0.17 [-0.72, 0.39] |
| Dambacher et al,2015 | 3.84 | 1.16 | 16 | 4 | 1.33 | 16 | 4.0% | -0.12 [-0.82, 0.57] |
| Conson et al,2015 | 616.69 | 150.4 | 16 | 627.82 | 151.56 | 16 | 4.0% | -0.07 [-0.77, 0.62] |
| Nord et al,2017 | 479.59 | 56.0674 | 31 | 477.19 | 53.3949 | 31 | 4.5% | 0.04 [-0.45, 0.54] |
| Qiao et al,2020 | 1,269.45 | 942.31 | 14 | 1,073.51 | 657.13 | 15 | 3.9% | 0.24 [-0.50, 0.97] |
| Brennan et al,2016 | 63.8 | 10.24 | 20 | 60.35 | 8.7 | 20 | 4.1% | 0.36 [-0.27, 0.98] |
| Okano et al,2013 | 751.4 | 71.5 | 10 | 723.7 | 45 | 10 | 3.4% | 0.44 [-0.45, 1.33] |
| Faehling and Plewnig,2016 | 1,170.09 | 45.57 | 21 | 1,110.98 | 34.62 | 22 | 4.0% | 1.44 [0.76, 2.12] |
| Fan et al,2020 | 2,374.777 | 437.793 | 18 | 1,448.777 | 124.477 | 18 | 3.3% | 2.81 [1.86, 3.76] |
| **Subtotal (95% CI)** | | | **225** | | | **227** | **46.8%** | **0.30 [-0.13, 0.73]** |
| Heterogeneity: Tau² = 0.44; Chi² = 52.46, df = 11 (P < 0.00001); I² = 79% | | | | | | | | |
| Test for overall effect: Z = 1.37 (P = 0.17) | | | | | | | | |
| **3.17.2 off RT** | | | | | | | | |
| Donaldson et al, 2019off | -618.944 | 236.218 | 17 | -57.727 | 72.658 | 18 | 3.1% | -3.18 [-4.21, -2.15] |
| Jafari et al,2021 2maoff | 427.16 | 46.45 | 13 | 569.58 | 45.11 | 14 | 2.8% | -3.02 [-4.17, -1.87] |
| Jafari et al,2021off | 495.08 | 42.27 | 14 | 569.58 | 45.11 | 14 | 3.5% | -1.65 [-2.53, -0.78] |
| Ayache et al, 2016off | 742.3 | 99.7 | 16 | 766.1 | 130.3 | 16 | 4.0% | -0.20 [-0.89, 0.49] |
| Clarke et al,2020 off | 972.21 | 50.33 | 79 | 979 | 50.13 | 79 | 4.8% | -0.13 [-0.45, 0.18] |
| Clarke et al,2020 benign | 949.68 | 52.14 | 79 | 955.82 | 52.04 | 79 | 4.8% | -0.12 [-0.43, 0.19] |
| Sanchez Lopez et al,2016R | 340 | 54 | 27 | 342 | 69 | 27 | 4.4% | -0.03 [-0.57, 0.50] |
| Sanchezlopez etal,2018of | 390 | 94 | 27 | 391 | 79 | 27 | 4.4% | -0.01 [-0.54, 0.52] |
| Boggio et al,2006 off | 209.15 | 77.62 | 26 | 205.45 | 61.62 | 26 | 4.3% | 0.05 [-0.49, 0.60] |
| Sagliano et al,2017off | 319.13 | 27.45 | 20 | 317.12 | 28.97 | 20 | 4.1% | 0.07 [-0.55, 0.69] |
| Marie Anne et al,2016off | 784.79 | 225.1 | 35 | 768.59 | 222.39 | 35 | 4.5% | 0.07 [-0.40, 0.54] |
| Kuehne et al,2019off | 723.05 | 72.29 | 18 | 700.9 | 58.84 | 18 | 4.0% | 0.33 [-0.33, 0.99] |
| Sanchez et al,2016off | 379.16 | 71.72 | 30 | 348.92 | 63.26 | 30 | 4.4% | 0.44 [-0.07, 0.95] |
| **Subtotal (95% CI)** | | | **401** | | | **403** | **53.2%** | **-0.41 [-0.80, -0.03]** |
| Heterogeneity: Tau² = 0.40; Chi² = 77.76, df = 12 (P < 0.00001); I² = 85% | | | | | | | | |
| Test for overall effect: Z = 2.09 (P = 0.04) | | | | | | | | |
| **Total (95% CI)** | | | **626** | | | **630** | **100.0%** | **-0.08 [-0.37, 0.21]** |
| Heterogeneity: Tau² = 0.42; Chi² = 138.59, df = 24 (P < 0.00001); I² = 83% | | | | | | | | |
| Test for overall effect: Z = 0.56 (P = 0.58) | | | | | | | | |
| Test for subgroup differences: Chi² = 5.85, df = 1 (P = 0.02), I² = 82.9% | | | | | | | | |

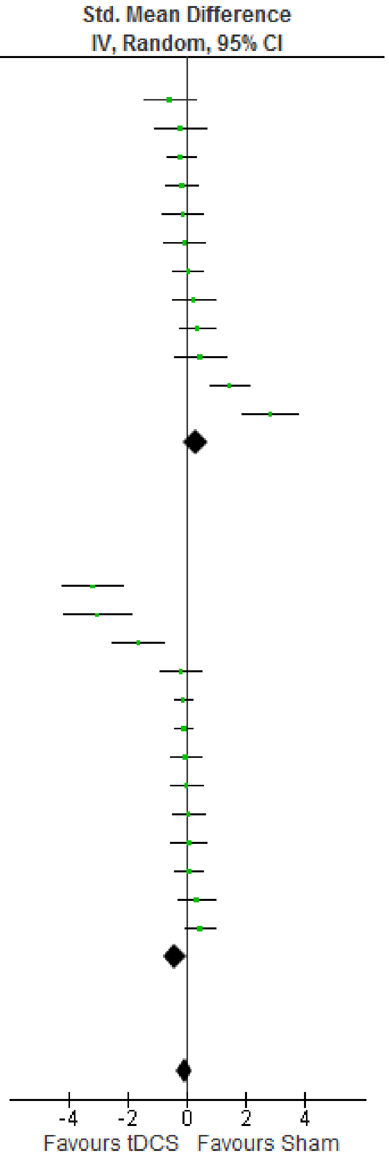


**Fig. S13** The subgroup meta-analysis based on the influence of online/offline of tDCS on emotion RT

| Study or Subgroup | tDCS Mean | tDCS SD | tDCS Total | Sham Mean | Sham SD | Sham Total | Weight | Std. Mean Difference IV, Random, 95% CI |
|---|---|---|---|---|---|---|---|---|
| **3.18.3 on ACC** | | | | | | | | |
| Faehling and Plewnig,2016 | 79.29 | 1.72 | 21 | 81.7 | 1.74 | 22 | 4.7% | -1.37 [-2.04, -0.70] |
| Nejati (d) et al,2021 | 3.42 | 0.92 | 22 | 4.05 | 1.29 | 22 | 4.9% | -0.55 [-1.16, 0.05] |
| Nejati et al,2021 | 2.69 | 2.69 | 16 | 3.95 | 2.91 | 16 | 4.6% | -0.44 [-1.14, 0.26] |
| Tremoliere et al,2018 | 94.12 | 16.35 | 34 | 97.06 | 11.94 | 34 | 5.2% | -0.20 [-0.68, 0.27] |
| Balzarotti&Colombo,2016 | 0.24 | 0.4116 | 14 | 0.25 | 0.3742 | 14 | 4.5% | -0.02 [-0.77, 0.72] |
| Nord et al,2017 | 92.54 | 5.011 | 31 | 91.5 | 5.6234 | 31 | 5.2% | 0.19 [-0.31, 0.69] |
| Qiao et al,2020 | 7.36 | 5.05 | 14 | 6.18 | 3.54 | 15 | 4.5% | 0.26 [-0.47, 1.00] |
| Brennan et al,2016 | 63.77 | 10.4648 | 20 | 60.35 | 8.5865 | 20 | 4.8% | 0.35 [-0.27, 0.98] |
| Schroeder et al,2015 | 0.96 | 0.0938 | 22 | 0.91 | 0.0938 | 22 | 4.9% | 0.52 [-0.08, 1.13] |
| Nejati(c) et al,2021 | 26.1 | 2.38 | 16 | 24.3 | 2.97 | 16 | 4.6% | 0.65 [-0.06, 1.37] |
| **Subtotal (95% CI)** | | | **210** | | | **212** | **47.9%** | **-0.06 [-0.42, 0.30]** |
| Heterogeneity: Tau² = 0.23; Chi² = 29.46, df = 9 (P = 0.0005); I² = 69% | | | | | | | | |
| Test for overall effect: Z = 0.33 (P = 0.74) | | | | | | | | |
| **3.18.4 off acc** | | | | | | | | |
| Feeser et al, 2013off | 0.26 | 0.27 | 48 | 0.56 | 0.36 | 48 | 5.4% | -0.94 [-1.36, -0.51] |
| Koenigs et al,2008off | 0.015 | 0.032 | 25 | 0.037 | 0.014 | 25 | 4.9% | -0.88 [-1.46, -0.29] |
| Weigand et al,2021off | 47.35 | 14.91 | 23 | 50.25 | 11.94 | 23 | 5.0% | -0.21 [-0.79, 0.37] |
| Gomez et al,2011off | 5.35 | 0.46 | 16 | 5.45 | 0.52 | 16 | 4.6% | -0.20 [-0.89, 0.50] |
| Ayache et al, 2016off | 88.4 | 15 | 16 | 90.2 | 14.6 | 16 | 4.6% | -0.12 [-0.81, 0.58] |
| Marie Anne et al,2016off | 88.25 | 6.69 | 35 | 86.67 | 7.03 | 35 | 5.2% | 0.23 [-0.24, 0.70] |
| Maeoka et al,2012off | 5.18 | 0.29 | 15 | 5.12 | 0.16 | 15 | 4.6% | 0.25 [-0.47, 0.97] |
| Boggio et al,2006 off | 10.67 | 7.05 | 26 | 8.71 | 2.98 | 26 | 5.0% | 0.36 [-0.19, 0.90] |
| Gilam et al, 2018off | 61 | 36.87 | 25 | 29.67 | 40.42 | 25 | 5.0% | 0.80 [0.22, 1.37] |
| Herrmann et al, 2016off | 0.12 | 0.56 | 14 | -0.49 | 0.32 | 12 | 4.2% | 1.27 [0.41, 2.13] |
| Donaldson et al, 2019off | 0.647 | 0.331 | 17 | -0.618 | 0.397 | 18 | 3.6% | 3.37 [2.30, 4.44] |
| **Subtotal (95% CI)** | | | **260** | | | **259** | **52.1%** | **0.29 [-0.25, 0.83]** |
| Heterogeneity: Tau² = 0.72; Chi² = 85.38, df = 10 (P < 0.00001); I² = 88% | | | | | | | | |
| Test for overall effect: Z = 1.04 (P = 0.30) | | | | | | | | |
| **Total (95% CI)** | | | **470** | | | **471** | **100.0%** | **0.11 [-0.22, 0.43]** |
| Heterogeneity: Tau² = 0.46; Chi² = 115.17, df = 20 (P < 0.00001); I² = 83% | | | | | | | | |
| Test for overall effect: Z = 0.65 (P = 0.52) | | | | | | | | |
| Test for subgroup differences: Chi² = 1.11, df = 1 (P = 0.29), I² = 9.6% | | | | | | | | |

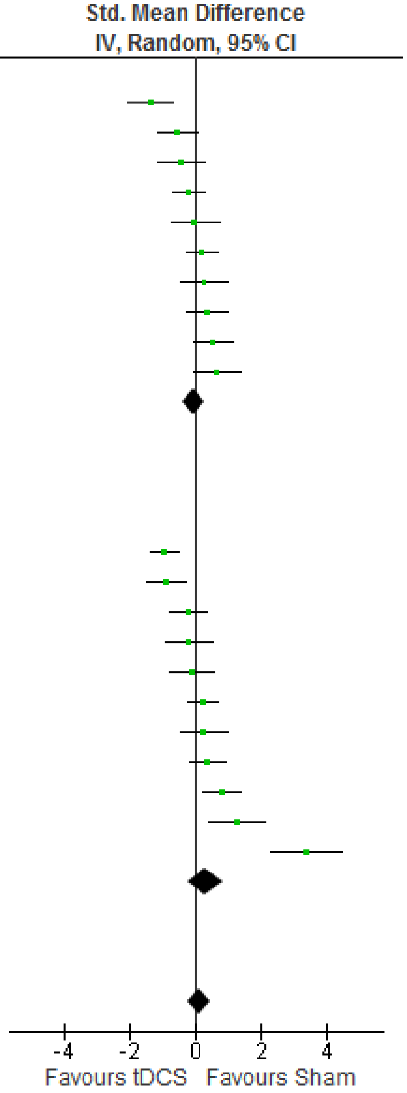


**Fig. S14** The subgroup meta-analysis based on the influence of online/offline of tDCS on emotion accuracy.

| Study or Subgroup | tDCS Mean | tDCS SD | tDCS Total | sham Mean | sham SD | sham Total | Weight | Std. Mean Difference IV, Random, 95% CI |
|---|---|---|---|---|---|---|---|---|
| **3.11.1 0-25years old** | | | | | | | | |
| Colombo et al,2016 | 4.18 | 3.1307 | 10 | 6.05 | 3.1307 | 10 | 3.9% | -0.57 [-1.47, 0.33] |
| Tremoliere et al,2018 | 13,000 | 6,392 | 34 | 14,693 | 9,599 | 34 | 5.0% | -0.21 [-0.68, 0.27] |
| Sarkar et al,2014 | 1,022.22 | 296.3 | 25 | 1,070.37 | 277.8 | 25 | 4.8% | -0.17 [-0.72, 0.39] |
| Dambacher et al,2015 | 3.84 | 1.16 | 16 | 4 | 1.33 | 16 | 4.5% | -0.12 [-0.82, 0.57] |
| Sanchez Lopez et al,2016R | 340 | 54 | 27 | 342 | 69 | 27 | 4.9% | -0.03 [-0.57, 0.50] |
| Sanchezlopez etal,2018of | 390 | 94 | 27 | 391 | 79 | 27 | 4.9% | -0.01 [-0.54, 0.52] |
| Marie Anne et al,2016off | 784.79 | 225.1 | 35 | 768.59 | 222.39 | 35 | 5.1% | 0.07 [-0.40, 0.54] |
| Qiao et al,2020 | 1,269.45 | 942.31 | 14 | 1,073.51 | 657.13 | 15 | 4.4% | 0.24 [-0.50, 0.97] |
| Kuehne et al,2019off | 723.05 | 72.29 | 18 | 700.9 | 58.84 | 18 | 4.6% | 0.33 [-0.33, 0.99] |
| Fan et al,2020 | 2,374.777 | 437.793 | 18 | 1,448.777 | 124.477 | 18 | 3.8% | 2.81 [1.86, 3.76] |
| **Subtotal (95% CI)** | | | **224** | | | **225** | **45.8%** | **0.17 [-0.22, 0.57]** |
| Heterogeneity: Tau² = 0.30; Chi² = 37.34, df = 9 (P < 0.0001); I² = 76% | | | | | | | | |
| Test for overall effect: Z = 0.86 (P = 0.39) | | | | | | | | |
| **3.11.2 >25years old** | | | | | | | | |
| Donaldson et al, 2019off | -618.944 | 236.218 | 17 | -57.727 | 72.658 | 18 | 3.5% | -3.18 [-4.21, -2.15] |
| Jafari et al,2021 2maoff | 427.16 | 46.45 | 13 | 569.58 | 45.11 | 14 | 3.2% | -3.02 [-4.17, -1.87] |
| Jafari et al,2021off | 495.08 | 42.27 | 14 | 569.58 | 45.11 | 14 | 4.0% | -1.65 [-2.53, -0.78] |
| Ayache et al, 2016off | 742.3 | 99.7 | 16 | 766.1 | 130.3 | 16 | 4.5% | -0.20 [-0.89, 0.49] |
| Clarke et al,2020 off | 972.21 | 50.33 | 79 | 979 | 50.13 | 79 | 5.4% | -0.13 [-0.45, 0.18] |
| Clarke et al,2020 benign | 949.68 | 52.14 | 79 | 955.82 | 52.04 | 79 | 5.4% | -0.12 [-0.43, 0.19] |
| Conson et al,2015 | 616.69 | 150.4 | 16 | 627.82 | 151.56 | 16 | 4.5% | -0.07 [-0.77, 0.62] |
| Nord et al,2017 | 479.59 | 56.0674 | 31 | 477.19 | 53.3949 | 31 | 5.0% | 0.04 [-0.45, 0.54] |
| Boggio et al,2006 off | 209.15 | 77.62 | 26 | 205.45 | 61.62 | 26 | 4.9% | 0.05 [-0.49, 0.60] |
| Sagliano et al,2017off | 319.13 | 27.45 | 20 | 317.12 | 28.97 | 20 | 4.7% | 0.07 [-0.55, 0.69] |
| Brennan et al,2016 | 63.8 | 10.24 | 20 | 60.35 | 8.7 | 20 | 4.7% | 0.36 [-0.27, 0.98] |
| Faehling and Plewnig,2016 | 1,170.09 | 45.57 | 21 | 1,110.98 | 34.62 | 22 | 4.5% | 1.44 [0.76, 2.12] |
| **Subtotal (95% CI)** | | | **352** | | | **355** | **54.2%** | **-0.42 [-0.89, 0.06]** |
| Heterogeneity: Tau² = 0.60; Chi² = 93.54, df = 11 (P < 0.00001); I² = 88% | | | | | | | | |
| Test for overall effect: Z = 1.70 (P = 0.09) | | | | | | | | |
| **Total (95% CI)** | | | **576** | | | **580** | **100.0%** | **-0.13 [-0.44, 0.19]** |
| Heterogeneity: Tau² = 0.45; Chi² = 133.90, df = 21 (P < 0.00001); I² = 84% | | | | | | | | |
| Test for overall effect: Z = 0.78 (P = 0.43) | | | | | | | | |
| Test for subgroup differences: Chi² = 3.45, df = 1 (P = 0.06), I² = 71.0% | | | | | | | | |

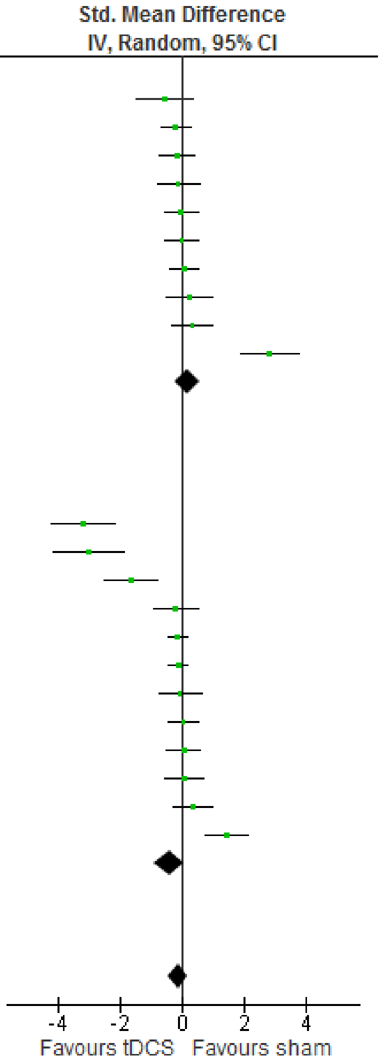


**Fig. S15** The subgroup meta-analysis based on the influence of age of tDCS on emotion RT.

| Study or Subgroup | tDCS Mean | tDCS SD | tDCS Total | sham Mean | sham SD | sham Total | Weight | Std. Mean Difference IV, Random, 95% CI |
|---|---|---|---|---|---|---|---|---|
| **3.12.1 0-25years old** | | | | | | | | |
| Nejati et al,2021 | 2.69 | 2.69 | 16 | 3.95 | 2.91 | 16 | 4.8% | -0.44 [-1.14, 0.26] |
| Weigand et al,2021off | 47.35 | 14.91 | 23 | 50.25 | 11.94 | 23 | 5.2% | -0.21 [-0.79, 0.37] |
| Tremoliere et al,2018 | 94.12 | 16.35 | 34 | 97.06 | 11.94 | 34 | 5.5% | -0.20 [-0.68, 0.27] |
| Gomez et al,2011off | 5.35 | 0.46 | 16 | 5.45 | 0.52 | 16 | 4.9% | -0.20 [-0.89, 0.50] |
| Balzarotti&Colombo,2016 | 0.24 | 0.4116 | 14 | 0.25 | 0.3742 | 14 | 4.7% | -0.02 [-0.77, 0.72] |
| Marie Anne et al,2016off | 88.25 | 6.69 | 35 | 86.67 | 7.03 | 35 | 5.5% | 0.23 [-0.24, 0.70] |
| Maeoka et al,2012off | 5.18 | 0.29 | 15 | 5.12 | 0.16 | 15 | 4.8% | 0.25 [-0.47, 0.97] |
| Qiao et al,2020 | 7.36 | 5.05 | 14 | 6.18 | 3.54 | 15 | 4.8% | 0.26 [-0.47, 1.00] |
| Nejati(c) et al,2021 | 26.1 | 2.38 | 16 | 24.3 | 2.97 | 16 | 4.8% | 0.65 [-0.06, 1.37] |
| Gilam et al, 2018off | 61 | 36.87 | 25 | 29.67 | 40.42 | 25 | 5.2% | 0.80 [0.22, 1.37] |
| **Subtotal (95% CI)** | | | **208** | | | **209** | **50.3%** | **0.11 [-0.14, 0.36]** |
| Heterogeneity: Tau² = 0.06; Chi² = 14.26, df = 9 (P = 0.11); I² = 37% | | | | | | | | |
| Test for overall effect: Z = 0.86 (P = 0.39) | | | | | | | | |
| **3.12.2 >25years old** | | | | | | | | |
| Faehling and Plewnig,2016 | 79.29 | 1.72 | 21 | 81.7 | 1.74 | 22 | 4.9% | -1.37 [-2.04, -0.70] |
| Feeser et al, 2013off | 0.26 | 0.27 | 48 | 0.56 | 0.36 | 48 | 5.7% | -0.94 [-1.36, -0.51] |
| Nejati (d) et al,2021 | 3.42 | 0.92 | 22 | 4.05 | 1.29 | 22 | 5.1% | -0.55 [-1.16, 0.05] |
| Ayache et al, 2016off | 88.4 | 15 | 16 | 90.2 | 14.6 | 16 | 4.9% | -0.12 [-0.81, 0.58] |
| Nord et al,2017 | 92.54 | 5.011 | 31 | 91.5 | 5.62 | 31 | 5.4% | 0.19 [-0.31, 0.69] |
| Brennan et al,2016 | 63.77 | 10.4648 | 20 | 60.35 | 8.5865 | 20 | 5.1% | 0.35 [-0.27, 0.98] |
| Boggio et al,2006 off | 10.67 | 7.05 | 26 | 8.71 | 2.98 | 26 | 5.3% | 0.36 [-0.19, 0.90] |
| Schroeder et al,2015 | 0.96 | 0.0938 | 22 | 0.91 | 0.0938 | 22 | 5.2% | 0.52 [-0.08, 1.13] |
| Herrmann et al, 2016off | 0.12 | 0.56 | 14 | -0.49 | 0.32 | 12 | 4.4% | 1.27 [0.41, 2.13] |
| Donaldson et al, 2019off | 0.647 | 0.331 | 17 | -0.618 | 0.397 | 18 | 3.7% | 3.37 [2.30, 4.44] |
| **Subtotal (95% CI)** | | | **237** | | | **237** | **49.7%** | **0.24 [-0.37, 0.86]** |
| Heterogeneity: Tau² = 0.88; Chi² = 90.96, df = 9 (P < 0.00001); I² = 90% | | | | | | | | |
| Test for overall effect: Z = 0.77 (P = 0.44) | | | | | | | | |
| **Total (95% CI)** | | | **445** | | | **446** | **100.0%** | **0.16 [-0.17, 0.48]** |
| Heterogeneity: Tau² = 0.45; Chi² = 106.31, df = 19 (P < 0.00001); I² = 82% | | | | | | | | |
| Test for overall effect: Z = 0.94 (P = 0.35) | | | | | | | | |
| Test for subgroup differences: Chi² = 0.16, df = 1 (P = 0.69), I² = 0% | | | | | | | | |

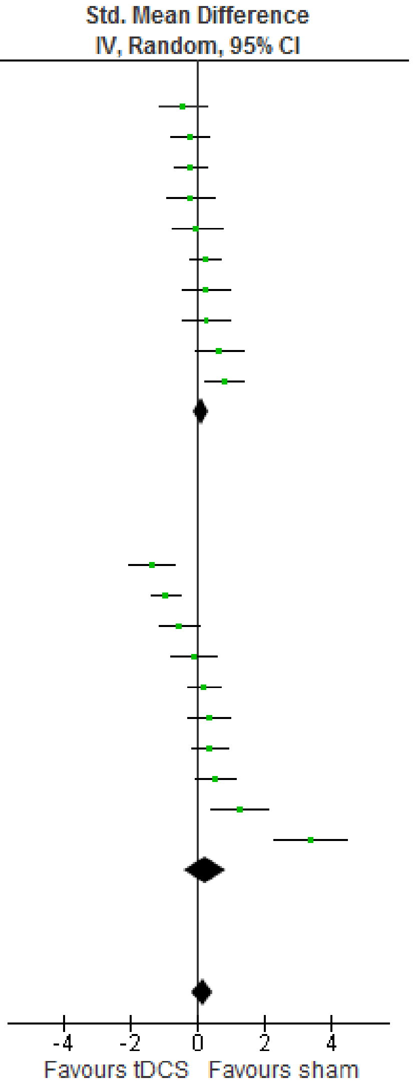


**Fig. S16** The subgroup meta-analysis based on the influence of age of tDCS on emotion accuracy.
